# In (deficit) schizophrenia, a general cognitive decline (G-CoDe) partly mediates the effects of neuro-immune and neuro-oxidative toxicity on the symptomatome and quality of life

**DOI:** 10.1101/2021.03.29.21254523

**Authors:** Michael Maes, Buranee Kanchanatawan

## Abstract

**Objective:** Schizophrenia and deficit schizophrenia are accompanied by neurocognitive impairments. The aim of this study was to examine whether a general factor underpins impairments in key Cambridge Neuropsychological Test Automated Battery (CANTAB) probes, verbal fluency (VFT), world list memory (WLM), true recall, and Mini Mental State Examination (MMSE).

**Methods:** We recruited 80 patients with schizophrenia and 40 healthy controls. All patients were assessed using CANTAB tests, namely paired-association learning (PAL), rapid visual information (RVP), spatial working memory (SWM), one touch stocking (OTS), intra/extradimensional set shifting (IED), and emotional recognition test (ERT).

**Results:** We found that a general factor, which is essentially unidimensional, underlies those CANTAB, VFT, WLM, True Recall, and MMSE scores. This common factor shows excellent psychometric properties and fits a reflective model and, therefore, reflects a general cognitive decline (G-CoDe) comprising deficits in semantic and episodic memory, recall, executive functions, strategy use, rule acquisition, visual sustained attention, attention set-shifting, and emotional recognition. Partial least Square analysis showed that 40.5% of the variance in G-Code is explained by CCL11, IgA to tryptophan catabolites, and increased oxidative toxicity; and that G-CoDe explains 44.8% of the variance in a general factor extracted from psychosis, hostility, excitation, mannerism, negative symptoms, formal thought disorders, and psychomotor retardation; and 40.9% in quality of life scores. The G-CoDe is significantly greater in deficit than in nondeficit schizophrenia.

**Conclusions:** A common core shared by a multitude of neurocognitive impairments (G-CoDe) mediates the effects of neurotoxic pathways on the phenome of (deficit) schizophrenia.

## INTRODUCTION

Recently, we have shown that the phenome of schizophrenia comprises three major components, namely a) the symptomatome comprising psychosis, hostility, excitation, mannerism, and negative (PHEMN) symptoms, psychomotor retardation (PMR), and formal thought disorders (FTD), b) the cognitome, namely the aggregate of cognitive dysfunctions, ^1^ and c) the phenomenome or the self-experiences of the illness as experienced by the patient. ^1^ Furthermore, we showed that a common general factor, which is essentially unidimensional, underlies PHEMN, PMR and FTD and, therefore, that this common core shared by those symptoms reflects overall severity of schizophrenia (OSOS). ^1-3^

Aberrations in the phenomenome consist of lowered self-rated, health-related quality of life (HR-QoL) with lowered scores on the physical health, psychological, social, and environmental health subdomains. ^1,4^ Aberrations in the cognitome comprise deficits in semantic and episodic memory, true recall, paired association learning, working memory, executive functions, set shifting, and attention. ^1,5-7^ Furthermore, these cognitome dysfunctions are strongly correlated with the symptomatome of schizophrenia and, therefore, may play a role in the onset and maintenance of the symptomatome and together may cause the lowered HR-QoL in that illness. ^1^

The aberrations in the cognitome, symptomatome, and phenomenome are more pronounced in deficit schizophrenia than in non-deficit schizophrenia. ^1,2,8^ The deficit syndrome or major neurocognitive psychosis is classically conceptualized as a subtype of schizophrenia which is characterized by the presence of negative symptoms with a significant functional impact on HR-QoL. ^4,7,9^ Nevertheless, we found that deficit schizophrenia is not only characterized by increased severity of negative but also by increased severity of PHEM symptoms, PMR and FTD. ^3,8,9^

The neurocognitive dysfunctions in (deficit) schizophrenia can adequately be assessed using cognitive tests, such as the Consortium to Establish a Registry for Alzheimer’s disease (CERAD) including the Mini mental State Examination (MMSE), ^9,10^ the Brief Assessment of Cognition in Schizophrenia (BACS), ^11^ and the computerized Cambridge Neuropsychological Test Automated Battery (CANTAB). ^5,9,12^ Nevertheless, it remains unknown whether the impairments in attention, semantic and episodic memory, working memory, executive functions, planning, and emotional recognition are distinct features of schizophrenia or whether they are intertwined manifestations of a general factor (or a single latent trait) reflecting a “general cognitive decline” (G-CoDe).

We also reported that a large part of the variance (47.6%) in a general factor extracted from PHEMN symptoms, PMR, FTD, and CERAD tests (semantic and episodic memory and recall), could mechanistically be explained by the effects of multiple neurotoxic pathways. ^7,13,14^ Thus, the latter authors reported that the cognitome and symptomatome of schizophrenia are strongly associated with a) increased oxidative stress toxicity (OSTOX) as indicated by a composite score comprising lipid hydroperoxides (LOOH), malondialdehyde (MDA), and advanced oxidation protein products (AOPP); b) lowered antioxidant defenses as indicated by a composite score comprising paraoxonase (PON)1 activity, sulfhydryl (-SH) groups, and total radical trapping parameter (TRAP); ^7^ c) increased levels of the neurotoxic and anti-neurogenesis chemokine C-C motif ligand 11 (CCL11) or eotaxin; ^15,16^ and d) increased levels of IgA levels directed to neurotoxic tryptophan catabolites (TRYCATs) including xanthurenic acid, picolinic acid, and 3-OH-kynurenine. ^6,17^ Activation of the TRYCAT pathway is not only significantly associated with deficits in world list memory, verbal fluency, and true recall as measured with the CERAD, but also with executive functions test scores as measured with CANTAB tests. ^6,15,16^

The mechanistic theory is that these different pathways have neurotoxic activities causing aberrations in gray and white matter functional plasticity including in neuronal circuits underpinning the cognitome and symptomatome of schizophrenia, such as “prefronto-temporal, prefronto-parietal, prefronto-striato-thalamic, and hippocampal and amygdalal neural circuits”. ^13,18-20^ Nevertheless, there are no data whether these biomarkers are associated with specific CANTAB test results or rather with G-CoDe as indicated by a general factor extracted from all CANTAB and CERAD tests.

Hence, the current study was conducted to examine a) whether in schizophrenia a general factor underpins different neurocognitive impairments as measured with CANTAB (paired association learning, rapid visual information processing, spatial working memory, intra/extradimensional set shifting, emotional recognition, executive functions) and CERAD (episodic and semantic memory, recall and MMSE) scores; and b) whether neurotoxic pathways (OSTOX, antioxidant defenses, CCL11, and IgA to TRYCATs) are associated with this general G-CoDe factor. Such results would indicate that a deficit in a common core shared by different neurocognitive impairments, which reflect dysfunctions in prefronto-temporal, prefronto-parietal, hippocampal, and fronto-striatal neural circuits, mediates the effects of neurotoxic immune and oxidative outcome pathways on the symptomatome and phenomenome of schizophrenia.

## Subjects and Methods

### Participants

This is a case-control study in Thai adults aged 18-65 years examining clinical and neuropsychological outcomes. We recruited schizophrenic outpatients who met the diagnostic criteria for schizophrenia of the DSM-IV-TR (and who were in a clinically stable state for at least one year) at the Department of Psychiatry, Faculty of Medicine, Chulalongkorn University, Bangkok, Thailand. The controls are healthy individuals recruited by word of mouth from the same catchment area. We excluded: 1) patients with a lifetime diagnosis of other axis-I DSM-IV / DSM-V mental disorders, including bipolar disorder, major depressive disorder, substance use disorders, and psycho-organic disorders; 2) healthy individuals with a lifetime diagnosis of any axis-I DSM-IV / DSM-V mental disorder and a family history of psychosis; and 3) healthy volunteers and schizophrenic patients with major medical illnesses, including rheumatoid arthritis, inflammatory bowel disease, multiple sclerosis, neurodegenerative disorders, chronic obstructive pulmonary disease, diabetes type 1 and 2, and alcohol use; patients who were treated with immunomodulatory drugs and antioxidant supplements; and c) pregnant and lactating women.

Primary deficit schizophrenia is defined according to criteria of the Schedule for the Deficit Syndrome (SDS): ^21^ at least 2 of the following 6 features must be present with a clinically significant severity: restricted affect, diminished emotional range, poverty of speech, curbing of interest, a diminished sense of purpose, and diminished social drive, for the preceding 12 months. Additionally, these symptoms should not be secondary to for example depression or extrapyramidal side effects induced by antipsychotic medication. All participants and their guardians (parents or other close family members) gave written informed consent prior to participation in this study. The study was conducted according to Thai and international ethics and privacy laws. Approval for the study was obtained from the Institutional Review Board of the Faculty of Medicine, Chulalongkorn University, Bangkok, Thailand (No 298/57), which is in compliance with the International Guideline for Human Research protection as required by the Declaration of Helsinki, The Belmont Report, CIOMS Guideline and International Conference on Harmonization in Good Clinical Practice (ICH-GCP).

### Methods

Both healthy individuals and schizophrenia patients underwent a comprehensive clinical interview performed by a senior research psychiatrist and a trained clinical research assistant with a master’s degree in mental health. We used a semi-structured interview, including the Mini-International Neuropsychiatric Interview (M.I.N.I.) in a validated Thai translation ^22^ and clinical assessments both made by the senior psychiatrist: 1) The Schedule for Deficit Syndrome (SDS); ^21^ 2) The Scale for the Assessment of Negative Symptoms (SANS); ^23^ 3) The Positive and Negative Syndrome Scale (PANSS); ^24^ 4) The Brief Psychiatric Rating Scale (BPRS); ^25^ and 5) the Hamilton Depression (HDRS). ^26^ We calculated z unit-weighted composite scores reflecting psychosis, hostility, excitation, mannerism, PMR, and FTD using different items of the BPRS, HDRS, and PANNS. ^13,14^ Electronic Supplementary File 1 (ESF 1), Table 1 **s**hows the items that were employed to compute these symptom subdomain scores. HR-QoL was assessed with the World Health Organization Quality of Life Instrument-Abbreviated version (WHO-QoL-BREF), ^27^ which measures four domains: 1) Domain 1 or physical health; 2) Domain 2 or psychological health; Domain 3 or social relationships; and 4) Domain 4 or environment. The raw scores were computed according to WHO-QoL-BREF criteria. ^27^ We also assessed tobacco use disorder (TUD) according to DSM-IV-TR criteria.

**Table 1.**
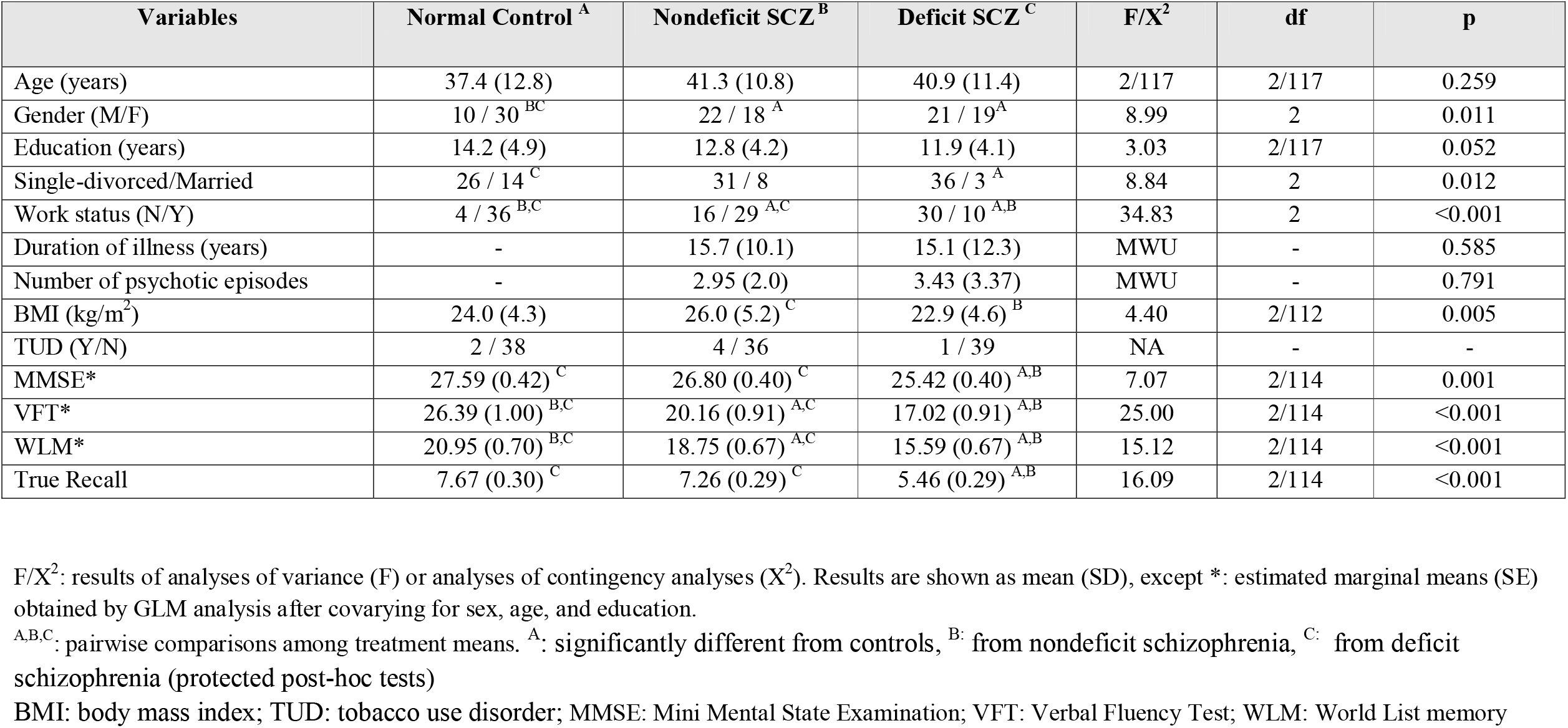
Demographic and clinical data in normal controls and schizophrenia patient with and without deficit syndrome.

| Variables | Normal Control <sup>A</sup> | Nondeficit SCZ <sup>B</sup> | Deficit SCZ <sup>C</sup> | F/X<sup>2</sup> | df | p |
| --- | --- | --- | --- | --- | --- | --- |
| Age (years) | 37.4 (12.8) | 41.3 (10.8) | 40.9 (11.4) | 2/117 | 2/117 | 0.259 |
| Gender (M/F) | 10 / 30 <sup>BC</sup> | 22 / 18 <sup>A</sup> | 21 / 19 <sup>A</sup> | 8.99 | 2 | 0.011 |
| Education (years) | 14.2 (4.9) | 12.8 (4.2) | 11.9 (4.1) | 3.03 | 2/117 | 0.052 |
| Single-divorced/Married | 26 / 14 <sup>C</sup> | 31 / 8 | 36 / 3 <sup>A</sup> | 8.84 | 2 | 0.012 |
| Work status (N/Y) | 4 / 36 <sup>BC</sup> | 16 / 29 <sup>AC</sup> | 30 / 10 <sup>AB</sup> | 34.83 | 2 | <0.001 |
| Duration of illness (years) | - | 15.7 (10.1) | 15.1 (12.3) | MWU | - | 0.585 |
| Number of psychotic episodes | - | 2.95 (2.0) | 3.43 (3.37) | MWU | - | 0.791 |
| BMI (kg/m<sup>2</sup>) | 24.0 (4.3) | 26.0 (5.2) <sup>C</sup> | 22.9 (4.6) <sup>B</sup> | 4.40 | 2/112 | 0.005 |
| TUD (Y/N) | 2 / 38 | 4 / 36 | 1 / 39 | NA | - | - |
| MMSE* | 27.59 (0.42) <sup>C</sup> | 26.80 (0.40) <sup>C</sup> | 25.42 (0.40) <sup>AB</sup> | 7.07 | 2/114 | 0.001 |
| VFT* | 26.39 (1.00) <sup>BC</sup> | 20.16 (0.91) <sup>AC</sup> | 17.02 (0.91) <sup>AB</sup> | 25.00 | 2/114 | <0.001 |
| WLM* | 20.95 (0.70) <sup>BC</sup> | 18.75 (0.67) <sup>AC</sup> | 15.59 (0.67) <sup>AB</sup> | 15.12 | 2/114 | <0.001 |
| True Recall | 7.67 (0.30) <sup>C</sup> | 7.26 (0.29) <sup>C</sup> | 5.46 (0.29) <sup>AB</sup> | 16.09 | 2/114 | <0.001 |

F/X<sup>2</sup>: results of analyses of variance (F) or analyses of contingency analyses (X<sup>2</sup>). Results are shown as mean (SD), except \*: estimated marginal means (SE) obtained by GLM analysis after covarying for sex, age, and education.
<sup>A,B,C</sup>: pairwise comparisons among treatment means. <sup>A</sup>: significantly different from controls, <sup>B</sup>: from nondeficit schizophrenia, <sup>C</sup>: from deficit schizophrenia (protected post-hoc tests)
BMI: body mass index; TUD: tobacco use disorder; MMSE: Mini Mental State Examination; VFT: Verbal Fluency Test; WLM: World List memory

### Cognitive assessments

Nine key Outcome Measures for CANTAB Research Suite Tests were used as advocated by CANTAB ^12^ to evaluate paired association learning (PAL), rapid visual information process test (RVP), spatial working memory (SWM), intra/extradimensional set shifting (IED), emotional recognition test (ERT), One touch Stockings of Cambridge (OTS). These CANTAB tests and the CERAD tests used in our study are described in ESF 1, CANTAB/CERAD tests.

### Assays

Blood was sampled at 8.00 a.m. after an overnight fast and thawed for assay. All assays were conducted in one and the same run by the same operator who was blinded to the clinical results. The assays are described in ESF 1, Assays.

### Statistics

We employed analysis of contingency tables (Χ^2^-test) to check associations between categorical variables and analysis of variance (ANOVA) or the Mann-Whitey-U test to assess differences in scale or ordinal variables between diagnostic classes (controls and schizophrenia with and without the deficit syndrome). We employed multivariate general linear model (GLM) analysis to assess the association between diagnostic groups and a set of CANTAB variables while adjusting for age, sex, education, and other background variables. When significant, we used tests for between-subject effects to check the associations between diagnosis and each of the CANTAB data. Results of multiple comparisons were always corrected for false discovery rate (FDR). ^29^ Subsequently, we computed model-generated estimated marginal mean (SE) values and employed protected, pairwise LSD tests to assess differences between the three diagnostic groups. Automatic (step-up) binary logistic regression analysis was employed to delineate the most important predictors of schizophrenia (versus controls) or deficit schizophrenia (versus non-deficit schizophrenia) using cognitive test results as discriminating variables. Power analysis showed that for an ANCOVA with three categories and five covariates, an effect size f = 0.30, power = 0.8, and α = 0.05, the required sample size should be around n=111. Given a possible drop-out rate of 8% we recruited 120 participants. Statistical analyses were performed using IBM SPSS Windows version 25. Tests were 2-tailed, and an alpha level of 0.05 indicated a statistically significant effect. All pattern recognition methods are described in ESF 1, Pattern Recognition Methods.

## Results

### Socio-demographic and clinical data

**Table 1** shows the demographic and clinical data for the three study groups. There were no significant differences in age and education between the three groups. In the schizophrenic study groups, there were significantly more men and single individuals without work than in the control group. There was no significant difference in duration of illness or number of psychotic episodes between deficit and nondeficit schizophrenia. BMI was significantly lower in patients with deficit schizophrenia than in those with nondeficit schizophrenia. Only seven people showed TUD. This table also shows the measurements of the three CERAD tests including MMSE and shows the estimated marginal means after adjusting for age, sex, and education years. The MMSE and True Recall scores were significantly lower in deficit schizophrenia than in the two other groups. VFT and WLM were significantly different between the three study groups.

ESF 2, Figure 1 shows that all symptom domain scores (except hostility) were significantly different between the three study groups and increased from controls to non-deficit and deficit schizophrenia. ESF 2, Figure 2 shows that the WHO-Qol domain scores were significantly different between the study groups and decreased from controls to non-deficit and deficit schizophrenia (except domain 3).

**Figure 1.**
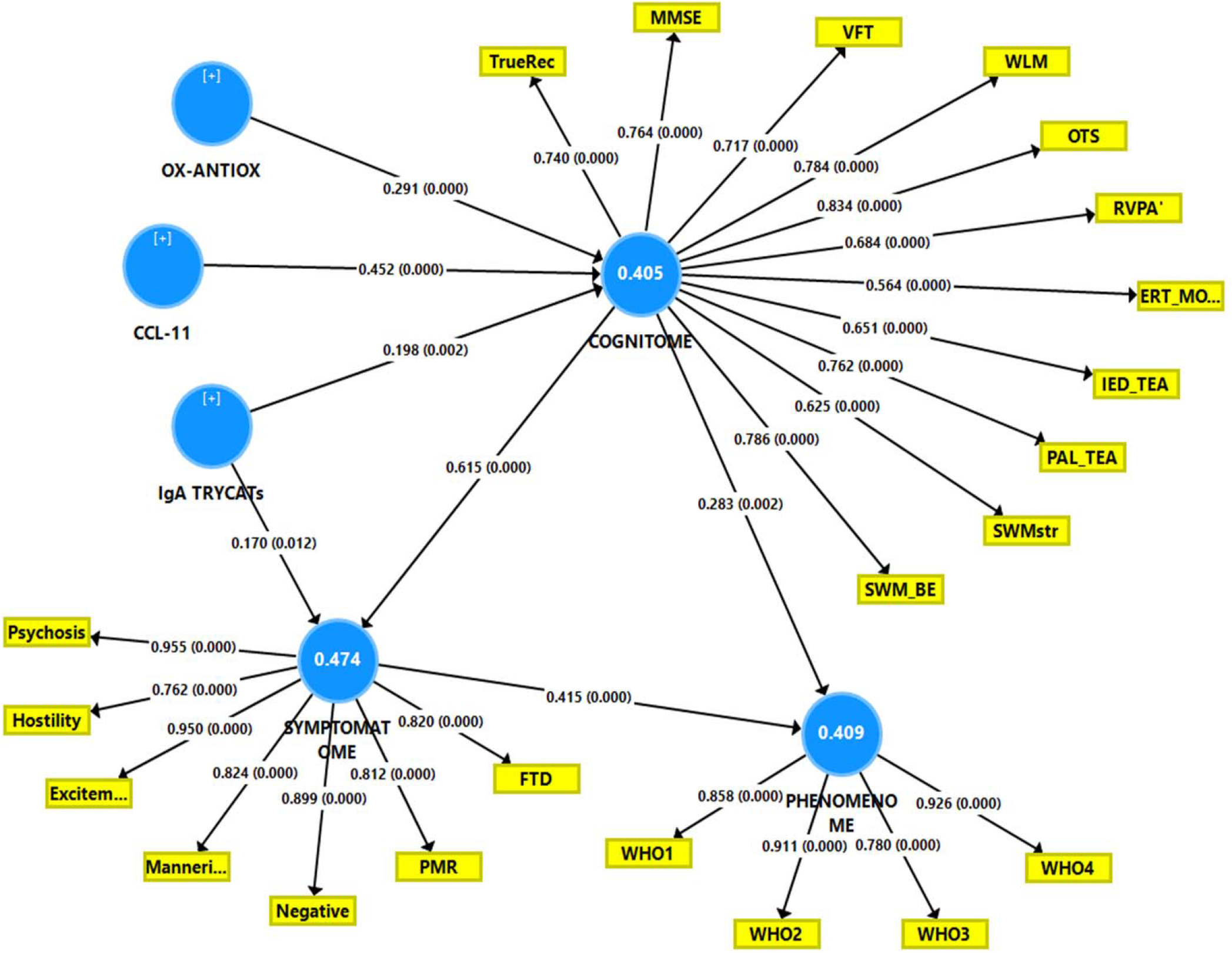
Results of Partial Least Squares analysis. OX-ANTIOX: ratio between oxidative stress toxicity/antioxidant defenses; IgA TRYCATs: IgA directed against tryptophan catabolites. TrueRec: True Recall; MMSE: Mini Mental State Examination; VFT: Verbal Fluency Test; WLM: World List memory. OTS: One touch stockings of Cambridge, problems solved on first choice; RVPA’: Rapid visual information process test, A’ prime; ERT_MORL: Emotional recognition test, median overall response latency; IED_TEA: Intra/extradimensional set shifting, total errors adjusted; PAL_TEA: Paired-association learning, total errors adjusted; SWM_STR: Spatial working memory, strategy; SWM_BE: Spatial working memory, between errors. Excitem: Excitement; Manneri: mannerism; Negative: negative symptoms; PMR: psychomotor retardation; FTD: formal thought disorders. WHO1-4: World Health Organization Quality of Life Instrument-Abbreviated version domains: 1) physical health; 2) psychological health; 3) social relationships; and 4) environment.

**Figure 2.**
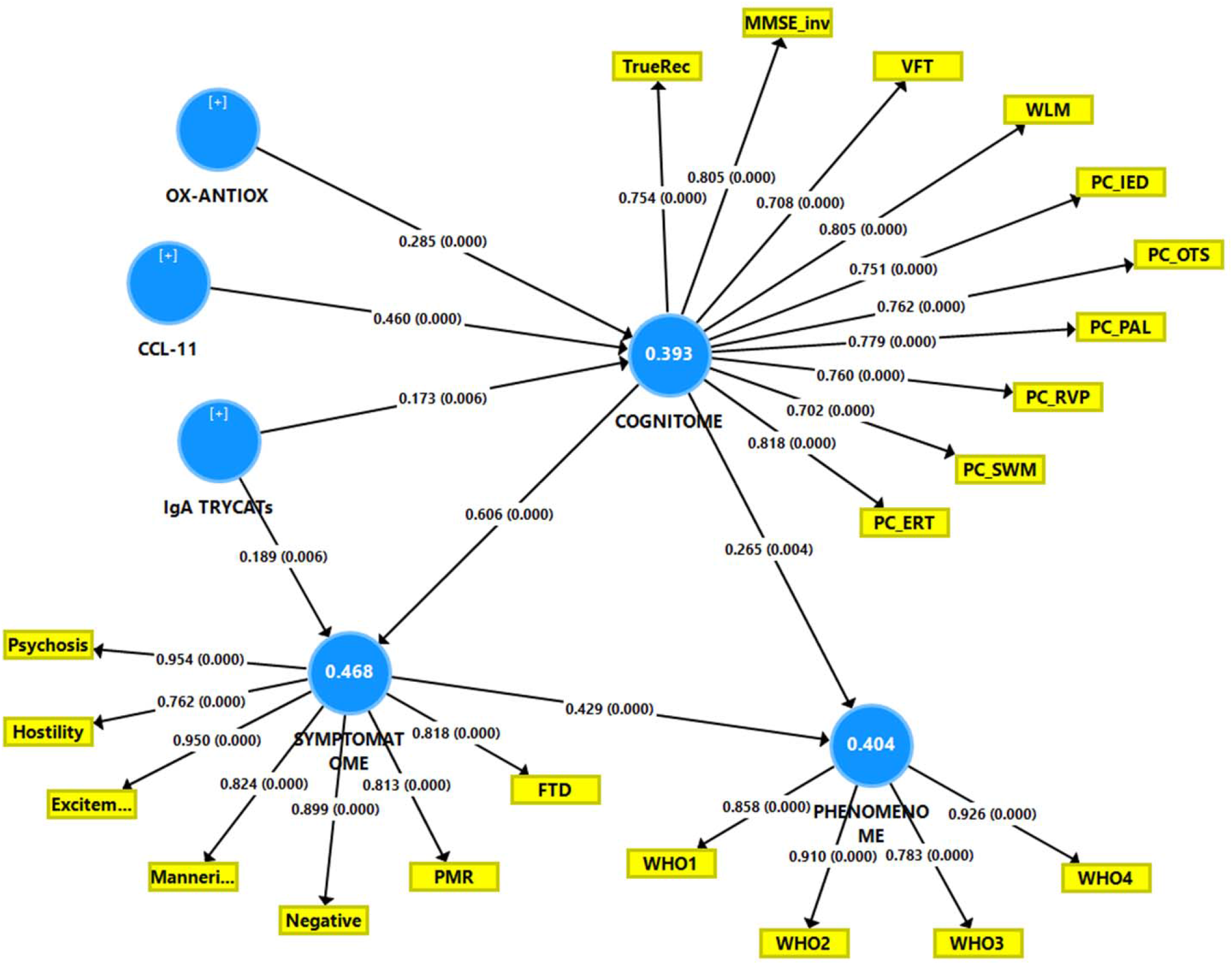
Results of Partial Least Squares analysis. OX-ANTIOX: ratio between oxidative stress toxicity/antioxidant defenses; IgA TRYCATs: IgA directed against tryptophan catabolites. TrueRec: True Recall; MMSE: Mini Mental State Examination; VFT: Verbal Fluency Test; WLM: World List memory. PC_: first principal component extracted from various test scores; IED: Intra/extradimensional set shifting tests; OTS: One touch stockings of Cambridge tests; PAL: Paired-association learning; RVP: Rapid visual information process; SWM: Spatial working memory; ERT: Emotional recognition test. Excitem: Excitement; Manneri: mannerism; Negative: negative symptoms; PMR: psychomotor retardation; FTD: formal thought disorders. WHO1-4: World Health Organization Quality of Life Instrument-Abbreviated version domains: 1) physical health; 2) psychological health; 3) social relationships; and 4) environment.

### CANTAB measurements in schizophrenia

**Table 2** shows the outcome of a multivariate GLM analysis which examined the associations between the diagnosis and the 9 CANTAB test results, while adjusting for age, education, and gender. We found a highly significant association between the diagnosis and the 9 CANTAB test results and significant effects of age and education, but not gender. Diagnosis shared around 23.1% of the variance with the CANTAB data. Univariate GLM analyses showed that there were significant associations between diagnosis and all CANTAB variables except IED_EDS and IED_TEA. False discovery rate correction did not change these results. **Table 3** shows the estimated marginal mean values of the 9 CANTAB measures in the three study groups. We found that participants with deficit schizophrenia showed worse outcomes on all measurements than controls (except IED_EDS). Moreover, PAL_TEA, RVP_A and ERT_MORL were significantly different between participants with deficit and nondeficit schizophrenia.

**Table 2.** Results of multivariate GLM analyses with the 10 CANTAB data as dependent variables and diagnosis as primary explanatory variable

| Test type | Dependent Variables | Explanatory Variables | F | df | p | Partial Eta Squared |
| --- | --- | --- | --- | --- | --- | --- |
| **Multivariate** | All 9 CANTAB | Diagnosis | 3.54 | 18/212 | <0.001 | 0.231 |
|  |  | Gender | 1.92 | 9/106 | 0.057 | 0.140 |
|  |  | Education | 6.41 | 9/106 | <0.001 | 0.352 |
|  |  | Age | 4.29 | 9/106 | <0.001 | 0.267 |
| **Univariate** | PAL_TEA | Diagnosis | 7.54 | 2/114 | 0.001 | 0.117 |
|  | RVP_A | Diagnosis | 6.04 | 2/110 | 0.003 | 0.099 |
|  | RVP_ML | Diagnosis | 6.84 | 2/114 | 0.002 | 0.107 |
|  | SWM_BE | Diagnosis | 17.63 | 2/114 | <0.001 | 0.236 |
|  | SWM_STR | Diagnosis | 16.48 | 2/113 | <0.001 | 0.137 |
|  | OTS_PSOFC | Diagnosis | 14.56 | 2/114 | <0.001 | 0.226 |
|  | IED_EDS | Diagnosis | 0.72 | 2/114 | 0.488 | 0.013 |
|  | IED_TEA | Diagnosis | 2.15 | 2/114 | 0.121 | 0.036 |
|  | ERT_MORL | Diagnosis | 8.56 | 2/114 | <0.001 | 0.131 |

Diagnosis: three groups, namely healthy controls, and patients with and without deficit schizophrenia
PAL\_TEA: Paired-association learning, total errors adjusted; RVP\_A: Rapid visual information process test, A' prime; RVP\_ML: Rapid visual information process test, median latency; SWM\_BE: Spatial working memory, between errors; SWM\_STR: Spatial working memory, strategy; OTS\_PSOFC: One touch stockings of Cambridge, problems solved on first choice; IED\_EDS: Intra/extradimensional set shifting, shift errors; IED\_TEA: Intra/extradimensional set shifting, total errors adjusted; ERT\_MORL: Emotional recognition test, median overall response latency

**Table 3.** Model-generated estimated marginal means (SE) obtained by univariate GLM analysis (age, sex and education adjusted)

| CANTAB Variables | Normal Controls <sup>A</sup> | Nondeficit SCZ <sup>B</sup> | Deficit SCZ <sup>C</sup> |
| --- | --- | --- | --- |
| PAL_TEA | 33.3 (8.4) <sup>C</sup> | 46.2 (7.9) <sup>C</sup> | 77.0 (8.0) <sup>A,B</sup> |
| RVP_A | 0.963 (0.010) <sup>C</sup> | 0.948 0.009 <sup>C</sup> | 0.917 0.009 <sup>A,B</sup> |
| RVP_ML | 377 (27) <sup>B,C</sup> | 484 (26) <sup>A</sup> | 513 (26) <sup>A</sup> |
| SWM_BE | 30.4 (3.4) <sup>B,C</sup> | 50.9 (3.2) <sup>A</sup> | 57.6 (3.2) <sup>A</sup> |
| SWM_STR | 34.2 (0.8) <sup>B,C</sup> | 38.7 (0.8) <sup>A</sup> | 40.7 (0.8) <sup>A</sup> |
| OTS_PSOFC | 8.27 (0.46) <sup>B,C</sup> | 5.94 (0.44) <sup>A</sup> | 4.80 (0.44) <sup>A</sup> |
| IED_EDS | 15.3 (2.0) | 18.0 (1.9) | 18.6 (2.0) |
| IED_TEA | 52.8 (7.4) <sup>C</sup> | 60.5 (7.0) | 73.8 (7.0) <sup>A</sup> |
| ERT_MORL | 2749 (248) <sup>B,C</sup> | 3525 (235) <sup>A,C</sup> | 4191 (237) <sup>A,B</sup> |

<sup>A,B,C</sup>: pairwise comparisons among treatment means. <sup>A</sup>: significantly different from controls, <sup>B</sup>: from nondeficit schizophrenia, <sup>C</sup>: from deficit schizophrenia (protected post-hoc tests)
PAL\_TEA: Paired-association learning, total errors adjusted; RVP\_A: Rapid visual information process test, A' prime; RVP\_ML: Rapid visual information process test, median latency; SWM\_BE: Spatial working memory, between errors; SWM\_STR: Spatial working memory, strategy; OTS\_PSOFC: One touch stockings of Cambridge, problems solved on first choice; IED\_EDS: Intra/extradimensional set shifting, shift errors; IED\_TEA: Intra/extradimensional set shifting, total errors adjusted; ERT\_MORL: Emotional recognition test, median overall response latency

ESF 2, confounding variables describes the effects of age, sex, and education on the CANTAB measurements. This section also describes that tobacco use disorder, BMI, employment, and the drug status did not have any significant effects on the CANTAB test results.

### Results of exploratory factor analysis

**Table 4** shows the results of an exploratory factor analysis with the CANTAB tests combined with the three CERAD tests and MMSE scores as variables. ESF 2, Exploratory factor analysis describes that one factor can be extracted from the listed CANTAB and CERAD tests and the MMSE.

**Table 4.** Results of exploratory factor analysis performed on key CANTAB and CERAD tests.

| Variables | Loadings | BCa bootstrap 95% confidence intervals |  |
| --- | --- | --- | --- |
| RVP_A' | **0.722** | 0.599 | 0.802 |
| SWM Strategy | **0.574** | 0.407 | 0.674 |
| PAL_TEA | **0.723** | 0.604 | 0.817 |
| RVP_ML | **0.571** | 0.393 | 0.684 |
| SWM_BE | **0.821** | 0.707 | 0.888 |
| OTS_PSOFC | **0.830** | 0.743 | 0.887 |
| IED_TEA | **0.579** | 0.438 | 0.700 |
| ERT_MORL | **0.526** | 0.351 | 0.680 |
| WLM | **0.703** | 0.578 | 0.816 |
| VFT | **0.663** | 0.515 | 0.753 |
| MMSE | **0.719** | 0.615 | 0.790 |
| True Recall | **0.641** | 0.518 | 0.770 |
| Kaiser-Meyer-Olkin test | 0.863 | 0.848 | 0.889 |
| Unidimensional Congruence | 0.965 | 0.955 | 0.979 |
| Explained Common Variance | 0.850 | 0.828 | 0.878 |
| Mean item residual absolute loadings | 0.232 | 0.203 | 0.246 |
| Goodness of Fit Index | 0.974 | 0.965 | 0.984 |
| Generalized H index | 0.923 | 0.922 | 0.960 |
| Weighted Root Mean Square Residual | 0.179 | 0.133 | 0.206 |

PAL\_TEA: Paired-association learning, total errors adjusted; RVP\_A: Rapid visual information process test, A' prime; RVP\_ML: Rapid visual information process test, median latency; SWM\_BE: Spatial working memory, between errors; SWM\_STR: Spatial working memory, strategy; OTS\_PSOFC: One touch stockings of Cambridge, problems solved on first choice; IED\_EDS: Intra/extradimensional set shifting, shift errors; IED\_TEA: Intra/extradimensional set
shifting, total errors adjusted; ERT\_MORL: Emotional recognition test, median overall response latency; MMSE: Mini Mental State Examination; VFT: Verbal Fluency Test; WLM: World List memory.

### Results of PLS analysis

ESF 2, PLS analysis shows the results of two PLS analyses and **Figure 1** and **Figure 2** show the outcome of those analyses. Consequently, we have computed the latent variable scores for the three CERAD scores (3CERAD), the three CERAD scores and the MMSE score (4CERAD), the 9 key CANTAB scores (9CANTAB), the 4 CERAD + 9 key CANTAB scores (CERAD+CANTAB), and the 7 PCs of the 7 CANTAB subdomains (PC7CANTAB). **Figure 3** shows a clustered bar graph of these 5 values in controls and schizophrenia patients with and without deficit syndrome. Univariate GLM analysis with age, sex, and education as covariates showed significant intergroup differences in the 3CERAD (F=29.54, df=2/110, p<0.001, partial eta squared=0.349), 4CERAD (F=29.05, df=2/110, p<0.001, partial eta squared=0.346), 9CANTAB (F=28.05, df=2/110, p<0.001, partial eta squared=0.285), CERAD+CANTAB (F=31.42, df=2/110, p<0.001, partial eta squared=0.364), and PC7CANTAB (F=16.39, df=2/110, p<0.001, partial eta squared=0.230). All scores were significantly different between the three groups and increased from controls → nondeficit → deficit schizophrenia. There was a strong association between CERAD+CANTAB and 3CERAD scores (r=0.834, p<0.001, n=116).

**Figure 3.**
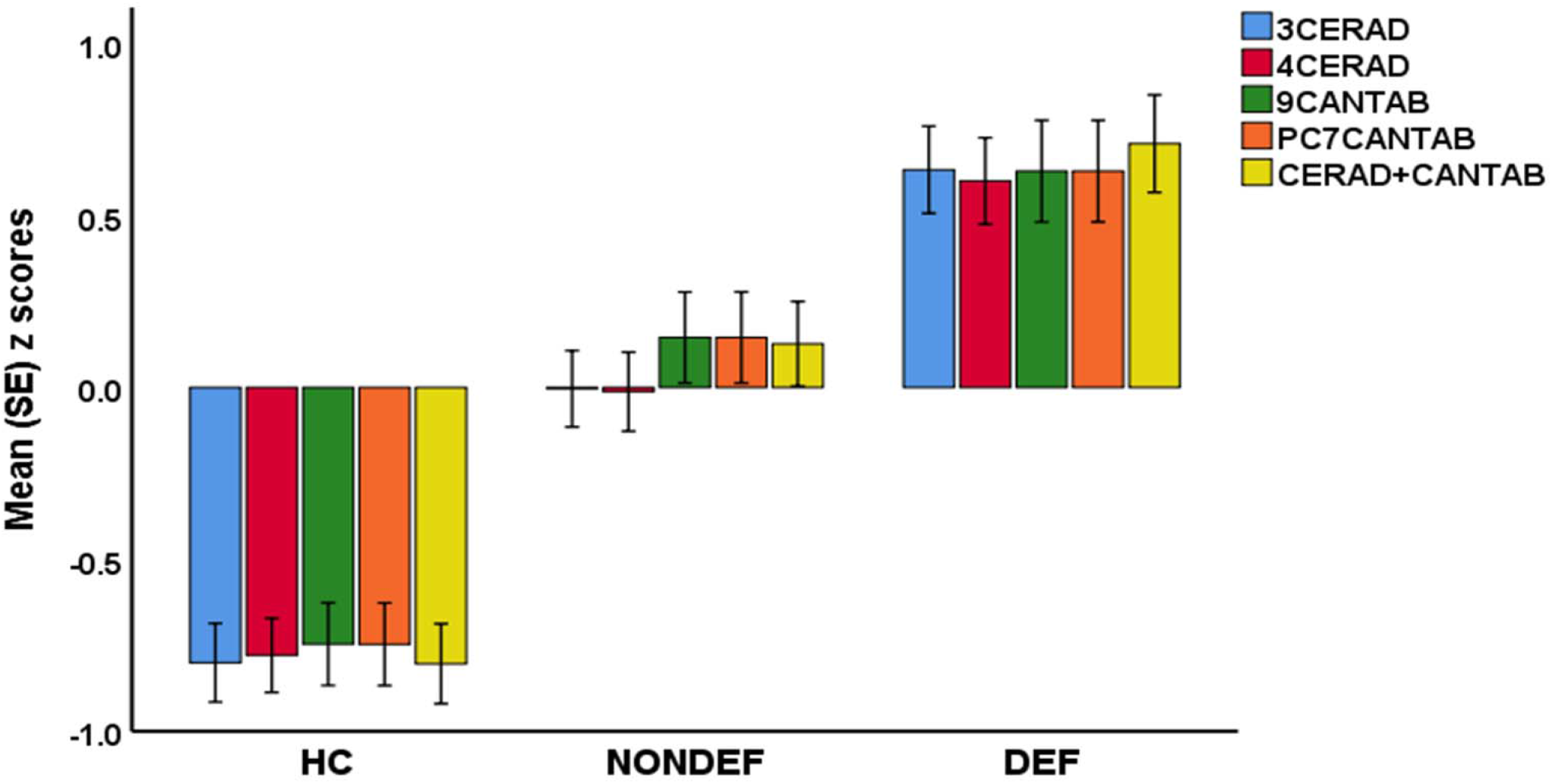
Clustered bar graph with neurocognitive scores in healthy controls (HC), and schizophrenia patients with (DEF) and without (NONDEF) deficit. 3CERAD: z unit-weighted composite score of Word List memory + Verbal Fluency + True Recall scores; 4CERAD: composite score of 3CERAD + MMSE (Mini Mental State Examination); 9CANTAB: composite score of 9 key CANTAB tests; PC7CANTAB: Principal component extracted from various test of 7 different domains; CERAD+CANTAB: computed as z unit-weighted composite score of 3 CERAD + 9CANTAB.

### Examination of all CANTAB tests

ESF 3, all CANTAB shows all CANTAB subtests that were measured in this study. ESF 3, Statistics describes how we extracted principal components (PC) from all CANTAB tests in the 7 CANTAB subdomains. ESF, Tables 1 and 2 show that there are important differences in these PCs extracted from all subdomain tests among the three study samples. ESF 3, Tables 3-5 show all the differences in all CANTAB subtests among the three study groups. ESF 3, Table 6 shows the results of binary logistic regression analyses discriminating schizophrenia from controls and deficit from non-deficit schizophrenia.

## Discussion

### The general cognitive decline (G-CoDe) in schizophrenia

The first major finding of this study is that, in schizophrenia, a general factor, which is essentially unidimensional, underpins the 9 key CANTAB, 3 CERAD (VFT, WLM, and True Recall) and MMSE scores. Moreover, this general factor fitted a reflective model and showed adequate psychometric properties indicating good convergent validity, excellent composite reliability, and adequate construct replicability. These findings indicate that, in schizophrenia, the aberrations in visual sustained attention, working memory and strategy use, rule acquisition and attention set-shifting, emotional recognition, semantic memory, episodic memory, and episodic memory recall are strongly interrelated and belong to one general factor, named G-CoDe. The G-CoDe factor accounts for 50.5% of the variance in the 12 cognitive scores and, therefore, summarizes the positive correlations among the impairments in all cognitive test scores in patients with schizophrenia. As such, the G-CoDe reflects aberrations in neuronal circuits, which underpin the deficits in CANTAB/CERAD tests and include prefronto-temporal, prefronto-parietal, hippocampal, and fronto-striatal neural circuits. ^2,13,18,19^

### The G-CoDe in nondeficit schizophrenia

The second major finding of this study is that nondeficit schizophrenia is characterized by significant impairments in visual memory, semantic memory, episodic memory recall, episodic memory and learning, spatial working memory and strategy use, spatial planning, rule acquisition and attention set-shifting, and interpretation of facial expression of emotion. As such, nondeficit schizophrenia is characterized by a general cognitive decline (namely the G-CoDe). Previously, Levaux et al. ^33^ reviewed CANTAB findings in schizophrenia and showed widespread cognitive impairments in working memory, decision-making, attention, visual memory, and especially in attentional set-shifting. Deficits in episodic memory, long-term memory, and conditional associative learning tasks are now well documented in schizophrenia. ^34,35^ Our findings on impaired visual memory in schizophrenia agree with those of Seidman et al. ^36^ and Kim et al. ^37^ who observed visual memory disorders in schizophrenia.

Moreover, our study also showed an overall decline in executive functions in schizophrenia, which agrees with the findings of Levaux et al. ^33^ and Pantelis et al. ^38^ The impaired SWM performance in schizophrenia suggests impaired working memory and strategy use, which extends previous findings. ^39-42^ Importantly, Bonner-Jackson et al. ^43^ reported that a deficit in the use of effective strategies in schizophrenia may modulate episodic memory performance. The current study showed significantly poorer IED scores in schizophrenia patients, suggesting impairments in rule acquisition and attention set-shifting, whereas no such results were described by Hilti et al. ^44^ Our OTS findings in schizophrenia patients indicate impairments in spatial planning, which agrees with previous reports. ^42,44,45^ Nevertheless, our factor analysis shows that the results on cognitive functioning in schizophrenia should not be discussed on a test-by-test basis, but rather by our new findings that all these test scores are features (reflective manifestations) of the G-CoDe.

### The G-CoDe in deficit schizophrenia

The third major finding of this study is that the severity of the G-CoDe, different key CANTAB tests, most PCs extracted from the subdomain tests scores, and many tests from the different CANTAB domains, and VFT, WLM, True Recall, and MMSE scores were significantly more pronounced in deficit than in nondeficit schizophrenia. Previous studies in deficit schizophrenia showed somewhat inconsistent results with significant or no significant changes in deficit versus nondeficit schizophrenia. ^46^ The latter review detected more impairments in general cognitive capacities and executive functions, sustained attention, and visuospatial memory in deficit than in nondeficit schizophrenia. Yu et al. ^46^ concluded that impairments in cognitive flexibility and sustained attention are the most important dysfunctions in deficit schizophrenia. But again, such deductions are less than optimal as deficit schizophrenia is characterized by increased severity of the general factor “G-CoDe”, which underpins the many cognitive features as assessed with CANTAB/CERAD.

### G-CoDe versus specificity of the cognitive CANTAB tests

Based on our findings that a general factor underpins all CANTAB/CERAD tests, we may conclude that it is not adequate to claim that the key CANTAB tests are specific tests, which reflect aberrations in distinct functions in schizophrenia. ^12^ If we would have employed only the key CANTAB tests we would have concluded that deficit schizophrenia is characterized by highly specific dysfunctions in paired-association learning, rapid visual processing, and interpretation of facial expression of emotion, whereas in fact schizophrenia is characterized by a G-CoDe in all CANTAB test scores. ESF 3, Discussion summarizes that many tests, which are not considered to be key tests by CANTAB, ^12^ are more relevant for (deficit) schizophrenia than the key tests of the same domain.

Finally, the three CERAD tests, VFT, WLM and True Recall, are more adequate to measure cognitive deficits in schizophrenia than the 9 key CANTAB tests, whilst a deficit in True Recall is the single best predictor of deficit from non-deficit schizophrenia. Moreover, the use of the CANTAB is hampered by the strong effects of education and age which explain a large part of the variability in the 9 key CANTAB tests, namely 35.2% and 26.7%, respectively. The WLM and True Recall tests results are much less prone to the effects of education and are not affected by age. ^17^ Consequently, we would propose that a composite score based on three CERAD tests, namely VFT, WLM, and True Recall, may be used as a convenient proxy for the G-CoDe in schizophrenia.

### The G-CoDe mediates the effects of neurotoxic pathways on the phenome of schizophrenia

The fourth major finding of this study is that the severity of the G-CoDe in schizophrenia is strongly predicted by neurotoxic pathways and partly mediates the effects of those pathways on the symptomatome (PHEMN, PMR, and FTD) and phenomenome (lowered HR-Qol scores). The severity of G-CoDe was to a large extent (40.2%) explained by increased oxidative stress toxicity and lowered antioxidant defenses (the OSTOX/ANTIOX ratio), CCL11, a neurotoxic chemokine that impacts neurogenesis, and increased activity of the TRYCAT pathway with increased IgA levels to the NOX/PRO ratio. ^14-17^ These results indicate that the neuro-immune and neuro-oxidative pathways impact G-CoDe through a multitude of neurotoxic effects including on neuroplasticity, synapse functions, protein regulation processes, cell signaling, apoptosis, and transcription. ^7,14^ As explained previously, these neurocognitive impairments may be causally associated with the PHEMN symptoms, FTD, and PMR of schizophrenia, and consequently with lowered HR-QoL. ^7,13-17^ Cognitive deficits are viewed as a direct result of “compromised cerebral function” ^47^ preceding psychotic symptoms. ^47,48^

It is heavily debated whether impairments in specific cognitive tests are associated with positive or negative symptoms. ^49-54^ For example, Lin et al. ^55^ found that negative symptoms may mediate the effects of cognition on functional outcomes. Yu et al. ^46^ reported significant correlations between negative symptoms and sustained attention, ideation fluency, cognitive flexibility, and visuospatial memory. Harvey ^48^ stated that the true nature of the associations between negative symptoms and cognitive impairments are still elusive. However, these discussions are not relevant, because PHEMN symptoms, FTD, and PMR are reflective manifestations of a general factor, namely OSOS. ^3,15^ Furthermore, the current study reported that the G-CoDe explained a large part (44.8%) of the variance in OSOS.

### Limitations

The results of the current study should be interpreted regarding its limitations. First, this is a case-control study, which does not allow to draw solid conclusions on causal models. Second, it would have been more interesting if we had performed functional neuroimaging techniques including multimodal imaging and imaging immunology ^56^ to detect which neuronal circuits are associated with the G-CoDe. Third, it could be argued why it was necessary to derive a general latent variable instead of using an existing observable variable such as the Global Assessment of Functioning (GAF) score. Nevertheless, until now, all studies in schizophrenia examined separate cognitive tests or a set of tests and no research has examined whether a general factor underpins all those neurocognitive deficits. Our results are important because they indicate that there are no specific disorders confined to one or more cognitive domains in schizophrenia, but rather a deficit in a common core shared by all neurocognitive score reflecting the influence of dysfunctions in prefronto-temporal, prefronto-parietal, hippocampal, and fronto-striatal neural circuits. In fact, the construction of our G-CoDe is comparable to the construction of the general factor g (general intelligence), which was conceptualized as a single general cognitive ability factor. ^57^ Moreover, we also showed that a combination of three CERAD tests (WLM, VFT, and True Recall) may be used as a proxy for the G-CoDe and that these CERAD tests perform better than the CANTAB tests. Importantly, our results show that rating scales, which measure the G-CoDe and OSOS in schizophrenia, should be based on reflective latent variable models and not on formative sum-scores. ^58^ Another strength of our research is that the current study employed the bottom-up, data-derived nomothetic network approach to delineate the associations among the building blocks of an illness, namely the adverse outcome pathways (neuro-immune and neuro-oxidative pathways), cognitome (the G-CoDe), symptomatome (the common core shared by PHEMN, PMR, and FTD, namely OSOS) and phenomenome (the common core shared by all domains of the HR-QoL). ^59,60,61^ This method allows to construct reliable and replicable nomothetic networks (our PLS models), which are generalizable disease models disclosing the causal links and mediating effects among the building blocks of the disorder. ^59,60^

### Conclusions

In schizophrenia, a general G-CoDe factor underlies impairments in semantic and episodic memory, recall, executive functions, strategy use, rule acquisition, visual sustained attention, attention set-shifting, and emotional recognition. Patients with deficit schizophrenia show a more profound deficit in this G-CoDe compared to those with nondeficit schizophrenia. The G-CoDe mediates the neurotoxic effects of immune and oxidative pathways on the symptomatome and phenomenome of schizophrenia.

## Supporting information

ESF 1

ESF 2

ESF 3

## Data Availability

The dataset generated during and/or analyzed during the current study will be available from the corresponding author upon reasonable request and once the dataset has been fully exploited by the authors.

## Compliance with ethical standards

### Disclosure of potential conflicts of interests

The authors report no conflict of interest with any commercial or other association in connection with the submitted article.

### Research involving human participants

Approval for the study was obtained from the Institutional Review Board of Chulalongkorn University, Bangkok, Thailand.

### Informed consent

All controls and patients and the guardians of patients gave written informed consent before participation in our study.

### Funding

This research has been supported by the Asahi Glass Foundation, Chulalongkorn University Centenary Academic Development Project.

### Author’s contributions

All the contributing authors have participated in the manuscript. MM and BK designed the study. BK recruited patients and completed diagnostic interviews and rating scales measurements. MM carried out the statistical analyses All authors contributed to interpretation of the data and writing of the manuscript.

## Notes

### Competing Interest Statement

The authors have declared no competing interest.

### Author Declarations

Institutional Review Board of the Faculty of Medicine, Chulalongkorn University, Bangkok, Thailand (No 298/57)

