## Supplementary material for "In (deficit) schizophrenia, a general cognitive decline (G-CoDe) partly mediates the effects of neuro-immune and neuro-oxidative toxicity on the symptomatome and quality of life": ESF 1

**ELECTRONIC SUPPLEMENTARY FILE 1 (ESF 1)**

https://scholar.google.co.th/citations?user=1wzMZ7UAAAAJ&hl=th&oi=ao

**ESF1, Table 1**. Indices of the different symptom domains and biomarker composite scores used in the current study

| **Symptom domains Biomarker scores** | **Z unit weighted composite scores** |
|---|---|
| Psychosis | sum of z score of item 1 on the positive subscale of the PANSS (zPANNSP1, delusion) *plus* zPANSSP3 (hallucinations) + zPANNSP6 (suspiciousness) *plus* z score of item 11 of the BPRS (zBPRS11: suspiciousness) *plus* zBPRS12 (hallucinatory behavior) *plus* zBPRS15 (unusual thought content). |
| Hostility | sum of zPANSSP7 (hostility) *plus* z-score of item 14 on the general psychopathology scale of the PANSS (zPANSSG14: poor impulse control) *plus* zBPRS10 (hostility) *plus* zBPRS14 (uncooperativeness). |
| Excitement | zPANNSP4 (excitement) *plus* zPANNSP5 (grandiosity) *plus* zBPRS8 (grandiosity) *plus* zBPRS17 (excitement). |
| Mannerism | zPANNSG5 *plus* zBPRS7 (both mannerism and posturing) |
| Formal thought disorders | zPANNSP2 (conceptual disorganization) *plus* item 5 of the PANNS negative subscale (PANNSN5: difficulty in abstract thinking) *plus* zBPRS4 (item 4 of the BPRS or conceptual disorganization) |
| Psychomotor retardation | z-score of HDRS item 8 (HDRS8: psychomotor retardation: slowness of thought and speech, decreased motor activity, impaired inability to concentrate) *plus* zPANSSG7 (reduction in motor activity as reflected in slowing or lessening of movements and speech, diminished responsiveness to stimuli and reduced body tone) *plus* zBPRS13 (reduction in energy level evidenced in slowed movements). |
| NOX/PRO TRYCAT | Ratio of noxious TRYCATs / generally more protective (NOX/PRO) TRYCATs computed as z score of IgA to picolinic acid + z IgA to xanthurenic acid + z IgA 3-OH-kynurenine – z IgA anthranilic acid – z kynurenic acid. |
| OSTOX/ANTIOX ratio | Ratio of oxidative stress toxicity / antioxidants computed as: z (z lipid hydroperoxides + z malondialdehyde + z advanced oxidation protein products) – z (z paraoxonase 1 activity + z sulphydryl groups + z total radical trapping parameter (TRAP) |

PANNS: Positive and Negative Syndrome Scale; BPRS: Brief Psychiatric Rating Scale (BPRS); HDRS: Hamilton Depression Rating Scale.

**ESF 1, CANTAB/CERAD tests**

a) Paired-association learning (PAL) to assess visual memory, episodic memory, and learning. We analyzed PAL total errors adjusted (PAL-TEA), "the number of times the subject chose the incorrect box for a stimulus on assessment problems but with an adjustment for the estimated number of errors they would have made on any problems, attempts & recalls they did not reach due to failing or aborting the test".

b) Rapid visual information process test (RVP) to assess visual sustained attention. We used RVP A' Prime (RVP_A), "the signal detection measure of sensitivity to the target, regardless of response tendency (the expected range is 0.00 to 1.00; bad to good). In essence, this metric is a measure of how good the subject is at detecting target sequences"; and RVP median latency (RVP_ML), "the median response latency during assessment sequence blocks where the subject responded correctly".

c) Spatial working memory (SWM) to assess working memory and strategy use. We analyzed SWM between errors (SWM-BE), "the total number of times the subject revisits a box in which a token has previously been found in the same problem"; and and SWM strategy (SWM-STR), for assessed problems with six boxes or more, the number of distinct boxes used by the subject to begin a new search for a token, within the same problem;

d) One touch stockings of Cambridge (OTS) to assess spatial planning. We analyzed OTS probability solved on first choice (OTS-PSOFC), "the number of assessment problems on which the first box choice made was correct".

e) Intra/extradimensional set shifting (IED) to assess rule acquisition and attention set-shifting. We measured IED EDS errors (IED_EDS), "the number of times that the subject failed to select the stimulus compatible with the current rule on the stage where the ED dimension shift occurs"; and IED total errors adjusted (IED-TEA), "the total number of times that the subject chose a stimulus incompatible with the current rule, plus, for each problem that was not reached (if any), an adjustment is made to the score";

f) Emotional recognition test (ERT), an emotional recognition task to recognize facial expression of emotion. We analyzed 2 ERT outcome measures, i.e. ERT mean overall response latency (ERT-MORL), "the median latency from stimulus onset to the subject's response button touch (i.e. emotion chosen) for all problems during assessment blocks. **ESF 2, Methods, Table 1** lists all CANTAB tests that were performed in the subjects.

We also measured three CERAD tests, [10] namely the Word List Memory (WLM) to assess verbal episodic memory, the Verbal Fluency Test (VFT) to assess semantic memory, and Word List True recall to assess verbal episodic memory recall. Finally, we also measured the Mini Mental State Examination (MMSE) to assess a more generalized cognitive deficit including in ideatoric and constructional praxis, orientation, speech, concentration, and memory.

***ESF 1, Assays.***

As described previously, [17] ELISA tests were employed to assay the plasma titers of immunoglobulins (Ig) A (IgA) directed against the tryptophan catabolites xanthurenic acid (XA) (Acros), picolinic acid (PA) (Acros), 3-OH-kynurenine (3HK) (Sigma), kynurenic acid (KA) (Acros), and anthranilic acid (AA) (Acros), linked to 20 mg BSA (ID Bio). Optical densities (ODs) were measured at 450 nm using Varioskan Flash (Thermo Scientific). Consequently, we computed the noxious / generally more protective (NOX/PRO) TRYCAT ratio as shown in ESF, Table 1**.** Serum CCL11 (R&D Systems, Inc, Minneapolis, MN, USA) was measured using the Bio-Plex® 200 System (Bio-Rad Laboratories, Inc.) as reported previously. [16] The methods for the OSTOX and ANTIOX assays were described previously: 7,28 "MDA levels were measured through complexation with two molecules of thiobarbituric acid using MDA estimation through high-performance liquid chromatography **(**HPLC Alliance e2695, Waters**'**, Barueri, SP, Brasil**)**. AOPP was quantified in a microplate reader (EnSpire, Perkin Elmer, USA) at a wavelength of 340 nm and is expressed in mM of equivalent chloramine T. LOOH was quantified by chemiluminescence in a Glomax Luminometer (TD 20/20), in the dark, at 30 °C for 60 min and the results are expressed in relative light units. TRAP was evaluated in a microplate reader (Victor X-3, Perkin Elmer, USA) and results are expressed in μM Trolox. -SH groups were evaluated in a microplate reader (EnSpire®, Perkin Elmer, USA) at a wavelength of 412 nm and results are expressed in μM. The substrates used to assess PON1 activity were phenyl acetate (PA, Sigma, USA) under high salt condition and CMPA (Sigma, USA), which is an alternative to the use of the toxic paraoxon. PON1 activities were determined by the rate of hydrolysis of CMPA (CMPAase) as well as by the rate hydrolysis of phenyl acetate under low salt condition. Analysis were conducted in a microplate reader (EnSpire, Perkin Elmer, USA). Consequently, we computed a z unit-weighted composite score reflecting the OSTOX/ANTIOX ratio as explained in ESF, Table 1.

***ESF 1, Pattern recognition methods***

Exploratory factor analysis (EFA), FACTOR, windows version 10.5.03 [30,31] was employed to explore the factor structure of the neurocognitive tests scores. The dispersion matrix used Pearson's correlations and we extracted factors with the robust unweighted least squares (RULS) method performed with 500 bias-corrected and accelerated (BCa) bootstraps. [30,31] The adequacy for factorization was checked with the Kaiser-Meyer-Olkin (KMO) test and Bartlett's test of sphericity. To estimate the number of factors to be retained we employed the Hull test and Parallel Analysis (Optimal Implementation) and Schwartz's Bayesian Information Criterion (BIC). We checked closeness to unidimensionality with explained common variance (ECV), unidimensional congruence (UNICO), and the mean of item residual absolute loadings (MIREAL). Data may be treated as essentially unidimensional when ECV >0.85, UNICO >0.95, and MIREAL <0.300. In order to check the model goodness-of-fit we employed the goodness-of-fit index (GFI). We assessed the distribution of residuals with the Weighted Root Mean Square Residual (WRMR) whereby values <1.0 indicate an adequate fit. We assessed construct replicability with the Generalized H index with values ≥0.80 indicating good stability across studies.

Subsequently, we conducted Partial Least Squares (PLS) path analysis (SmartPLS), [32] which was used to examine the associations between a latent vector (LV) extracted from the 9 key CANTAB, MMSE, VFT, WLM, and True Recall scores (reflecting G-CoDe) in a reflective model, and the biomarkers (entered as single indicators), the symptomatome (a LV extracted from PHEMN symptoms, FTD and PMR) and the phenomenome (a LV extracted from the 4 WHO-QoL domains). The three single indicators, namely the OSTOX/ANTIOX ratio, CCL11, and IgA NOX/PRO, predicted the cognitome, symptomatome, and phenomenome. We conducted complete PLS path analysis only when the constructs and the model fit complied with pre-specified quality criteria, namely: a) all factor loadings are significant ($p<0.001$) and > 0.500 (by preference 0.660); b) the LVs show good construct validity or internal consistency reliability and convergent validity as indicated by Cronbach alpha > 0.750, composite reliability > 0.800, rho_A >0.800, and average variance extracted (AVE) > 0.500; c) the model fit is adequate as assessed with the standardized root mean residual (SRMR) < 0.08; and d) the cross-validated predictive relevance of the PLS path model is adequate as assessed with the cross-validated redundancy approach with blindfolding, a predictive sample re-use technique. The latter was performed with an omission distance of 7 to compute the Stone-Geisser's $Q^2$ statistic whereby values of $Q^2 > 0.02$, 0.15, and 0.35 indicate that the model has a small, medium, and large predictive relevance for the selected construct. Indicators of the inner and outer model or constructs that do not comply with those quality data are eliminated from the final model. Subsequently, complete PLS bootstrapping (with 5000 bootstrap samples) is performed with calculation of the t-values and loadings of the indicators in the outer model, and the path coefficients for the inner model. Total, total indirect, and specific indirect effects are calculated. Confirmatory Tetrad Analysis (CTA) is conducted to assess possible misspecifications of a reflective LV model.
