## Supplementary material for "In (deficit) schizophrenia, a general cognitive decline (G-CoDe) partly mediates the effects of neuro-immune and neuro-oxidative toxicity on the symptomatome and quality of life": ESF 2

**ELECTRONIC SUPPLEMENTARY FILE 2 (ESF 2)**

https://scholar.google.co.th/citations?user=1wzMZ7UAAAAJ&hl=th&oi=ao

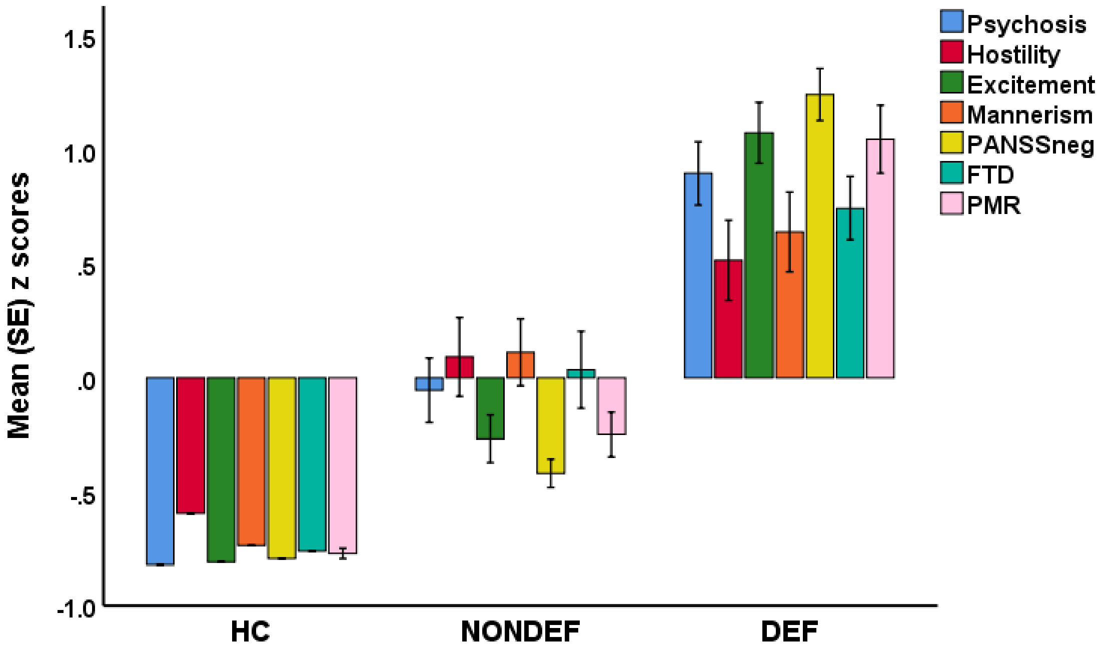


***ESF 2, Figure 1*. Clustered bar graph showing the symptomatome domains of schizophrenia.**
**in healthy controls (HC) and patients with nondeficit (NONDEF) and deficit (DEF) schizophrenia.**
PANNSneg: the negative subscale of the Positive and Negative Syndrome Scale; FTD: formal thought disorders; PMR: psychomotor retardation.
Multivariate GLM analysis adjusted for age, sex, and education shows a significant association between diagnosis and the symptom domains (F=22.13, df=14/214, p<0.001, partial eta squared=0.602). Pairwise comparisons among treatment means showed that all domain scores were significantly different between the three classes (except hostility which did not differ between both schizophrenia groups).

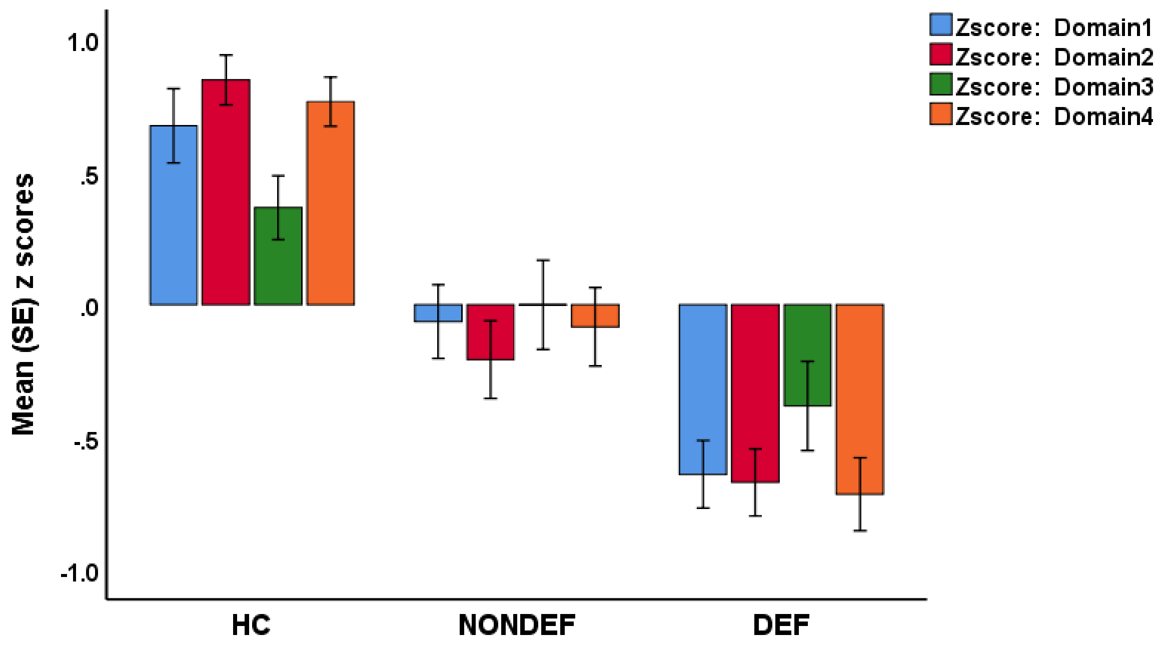


***ESF 2, Figure 2.*** **Clustered bar graph showing the World Health Organization Quality of Life scores (in z values) in healthy controls (HC) and patients with nondeficit (NONDEF) and deficit (DEF) schizophrenia.**

Shown are the mean z scores of domains 1: physical health; domain 2: psychological health; domain 3: social relationships; and domain 4: environment.

Multivariate GLM analysis adjusted for age, sex, and education shows a significant association between diagnosis and the WHO-QoL domains ($F=9.80$, $df=8/218$, $p<0.001$, partial eta squared=0.265). Pairwise comparisons among treatment means showed that all domain scores were significantly different between the three classes, except domain 3 which was different between HC and DEF only.

***ESF 2, confounding variables*.**

Univariate GLM analyses (with diagnosis, sex and education as additional explanatory variables) and false discovery rate p correction showed that age was significantly and positively associated with SWM_BE (p=0.005), SWM_STR (p=0.007) and ERT_MORL (p=0.042) and significantly and negatively with OTS_PSOFC (p=0.005). Univariate GLM analyses (with diagnosis, sex, and age as additional explanatory variables) and false discovery rate p correction showed that years of education was significantly and positively associated with RVP_A (p=0.002), OTS_PSOFC (p=0.002) and negatively with PAL_TEA (p=0.015), SWM_BE (p=0.015), and IED_TEA (p=0.002). Tobacco use disorder (F=1.86, df=10/100, p=0.060), BMI (F=1.01, df=10/979, p=0.442) and employment status (F=0.90, df=10/100, p=0.535) did not show a significant effect upon CANTAB performance, while the effects of diagnosis remained significant.

The effects of drug treatment of the participants were also investigated. Prescribed medications were risperidone: n=32, olanzapine: n=5, quetiapine n=4, clozapine: n=10, haloperidol n=10, perphenazine n=19, chlorpromazine n=4, fluphenazine n=10, antidepressants n=28, mood stabilizers n=12, anxiolytics n=29, and trihexyphenidyl n=47. Using univariate GLM analysis no significant effects of these drugs (or their dosages) could be found on any of the 9 CANTAB or 4 CERAD tests.

***ESF 2, Exploratory factor analysis***

**Table 4** shows the results of an exploratory factor analysis with the CANTAB tests combined with the three CERAD tests and MMSE scores as variables. A first analysis showed that all variables had loadings > 0.500, except IED_EDS (0.103) and, therefore, we have eliminated the latter CANTAB test score from the final analysis shown in Table 4. The KMO statistic of sampling adequacy was 0.863 and Bartlett's test ($\chi 2=810.0$, df=66, $p<0.00001$) show adequate factorability of the correlation matrix. We found that one real-data eigenvalue was greater than 1.0, and that the first factor explained 50.5% of the variance in the 12 cognitive scores. The BIC and Hull test, and PA analysis indicated that the advised number of factors was one. Moreover, the ECV (≥0.850), UNICO (≥0.95), and MIREAL (≤0.3) values indicated that the first factor extracted from the 12 cognitive tests should be treated as unidimensional. Table 4 shows that all cognitive scores loaded highly (> 0.500) and that the model goodness of fit index showed adequate values. In addition, also the distribution of residuals as assessed with weight root mean square residuals showed a good fit. Finally, the Generalized H index of 0.923 indicated an adequate construct replicability and performance across studies.

***ESF 2, PLS analysis***.

**Figure 1** shows the results of a first PLS analysis with a LV extracted from the four HR-QoL subdomain scores as final output variable, and with the symptomatome (introduced as a latent vector extracted from PHEMN symptoms, PMR, and FTD) and the cognitome (latent vector extracted from the key CANTAB tests, MMSE, VFT, WLM, and True Recall) as direct input variables, which may mediate the effects of the biomarkers which were entered as single indicators, namely IgA NOX/PRO TRYCATs, CCL11, and the OSTOX/ANTIOX ratio.

The model fit was more than adequate with a SRMR of 0.052 for the saturated model and 0.054 for the estimated model. The construct validities of all latent vectors were adequate with Cronbach α > 0.893, composite reliabilities > 0.923, rho_A > 0.911, and AVEs > 0.523. The loadings on all LVs were > 0.500 at p<0.001. The cognitome LV showed a Cronbach α of 0.907 and an AVE of 0.523 with all loadings > 0.651, except ERT_MORL (0.564). PLS path analysis conducted with 5000 bootstraps showed that 40.9% of the variance in the phenomenome LV was explained by the symptomatome and cognitome LV; 47.4% of the variance in the symptomatome LV was explained by the cognitome and IgA TRYCATs; and 40.5% of the variance in the cognitome was explained by the cumulative effects of the OSTOX/ANTIOX ratio, IgA TRYCATS, and CCL11. We found that all the mediated effects of the three biomarkers on the symptomatome and phenomenome were significant. The cognitome (see Figure 1) has significant direct and specific indirect effects on the phenomenome mediated by the symptomatome (t=4.06, p<0.001). Blindfolding showed that the replicability of the cognitome LV was adequate, namely cross-validated redundancy = 0.203. Multi-group analysis showed that there were no significant differences between both sexes in the paths, indirect and direct effects. Confirmatory Tetrad Analysis showed that the cognitome LV was not misspecified as a reflective model.

Consequently, we have performed a second PLS analysis which is quite similar to the analysis shown in Figure 1 except that we used some other indicators for the cognitome LV, namely the first PCs extracted from all IED, OTS, PAL, RVP, SWM, and ERT tests, and in addition the three CERAD tests including MMSE. The model fit was more than adequate with a SRMR of 0.050 for the saturated model and 0.053 for the estimated model. The construct validity of the cognitome LV was particularly good with Cronbach $\alpha = 0.921$, composite reliability = 0.934, rho_A = 0.924, and AVEs > 0.586. **Figure 2** shows that the loadings on this LV were > 0.702 at $p<0.001$. Figure 2 shows that 39.9% of the variance in the cognitome was explained by the three biomarkers. We found that the three biomarkers had significant effects on the symptomatome and the phenomenome, which were mediated by the cognitome. The latter showed significant direct (see Figure 2) and specific indirect ($t=4.31$, $p<0.001$) effects on the phenomenome, and that the latter were mediated by the symptomatome.
