## Supplementary material for "In (deficit) schizophrenia, a general cognitive decline (G-CoDe) partly mediates the effects of neuro-immune and neuro-oxidative toxicity on the symptomatome and quality of life": ESF 3

**ELECTRONIC SUPPLEMENTARY FILE 3 (ESF 3)**

https://scholar.google.co.th/citations?user=1wzMZ7UAAAAJ&hl=th&oi=ao

***ESF 3, all CANTAB.***

a) Motor screening test (MOT). We analyzed 4 MOT variables, i.e. MOT mean latency (MOT-ML), MOT mean error (MOT-ME), MOT total correct (MOT-TC) and MOT total errors (MOT-TE).

b) Paired-association learning (PAL). We analyzed 11 PAL variables, i.e. PAL first trial memory score (PAL-FTMS), PAL mean errors to success (PAL-METS), PAL mean trials to success (PAL-MTTS), PAL number of patterns reached (PAL-NPR), PAL number of patterns succeeded (PAL-NPS), PAL stages completed (PAL-SC), PAL stages completed on first trial (PAL-SCFT), PAL total errors (PAL-TE), PAL total errors adjusted (PAL-TEA), PAL total trials (PAL-TT) and PAL total trials adjusted (PAL-TTA).

c) One touch stockings of Cambridge (OTS). We analyzed 6 OTS outcome measures, i.e. OTS mean latency to correct (OTS-MLTC), OTS mean latency to first choice (OTS-MLTFC), OTS mean choices to correct (OTS-MCTC), OTS probability of error given correct (OTS-PEGC), OTS probability of error given error (OTS-PEGE) and OTS probability solved on first choice (OTS-PSOFC).

d) Rapid visual information process test (RVP). We used 11 RVP outcome measures, i.e. RVP A, RVP probability of false alarm (RVP-PFA), RVP probability of hit (RVP-PH), RVP probability of hit blocks 1-7 (RVP-PHB1-7), RVP mean latency (RVP-ML), RVP total correct rejections (RVP-TCR), RVP total false alarms (RVP-TFA), RVP total hits (RVP-TH), RVP total hits blocks 1-7 (RVP-THB1-7), RVP total misses (RVP-TM), RVP total misses blocks 1-7 (RVP-TMB1-7).

e) Spatial working memory (SWM). We analyzed 9 SWM outcome measures, i.e. SWM between errors (SWM-BE), SWM between errors 4 boxes (SWM-BE4B), SWM strategy (SWM-STR), SWM strategy 4-10 boxes (SWM-STR4-10B), SWM mean time to first response (SWM-MTFR), SWM mean time to last response (SWM-MTLR), SWM mean token preparation time (SWM-MTPT), SWM total errors (SWM-TE) and SWM total error 4 boxes (SWM-TE4B).

f) Intra/extradimensional set shifting (IED). We assessed 10 outcome measures, i.e. IED completed stage errors (IED-CSE), IED complete stage trials (EID-CST), IED errors block 1 (IED-EB1), IED Pre-ED errors (IED-PEE), IED stages completed (IED-SC), IED total errors (IED-TE), IED total errors adjusted (IED-TEA), IED total latency (IED-TL), IED total trials (IED-TT) and IED total trials adjusted (IED-TTA).

g) Emotional recognition test (ERT). We analyzed 3 ERT outcome measures, i.e. ERT mean overall response latency (ERT-MORL). ERT percent correct (ERT-PC), and ERT total number correct (ERT-TNC).

***ESF 3, Statistics***

We used factor analyses, principal component (PC) method (using SPSS25) to reduce the number of CANTAB variables, namely 4 MOT, 11 PAL, 11 RVP, 9 SWM, 6 OTS, 10 IED, and 3 ERT tests into 7 interpretable PCs reflecting the same subdomains. Towards this end we have extracted the first PC of each CANTAB subtest. The first PC of the 4 MOT tests explained 65.6% of the variance, the first PC in the PAL data explained 76.6% of the variance, RVP: 76.8%, SWM: 64.5%, OTS: 68.5%, IED: 48.7% and ERT: 75.3%. As such we have delineated seven interpretable factors, which reflect the total variability in the CANTAB subdomains. Subsequently, we have entered the seven first PC scores in multivariate GLM analysis. If the overall GLM analysis performed on the 7 PCs showed significant effects of diagnosis and the tests of between-subject effects on a particular PC were significant, we also examined the effects of diagnosis on all CANTAB tests comprising that subdomain. We also examined whether a latent trait could be extracted from these 7 PCs and found that a first PC explained 57.2% of the variance in the 7 CANTAB tests which were all highly loaded on this first PC (all > 0.750, except PC MOT which showed a loading of 0.539).

***ESF 3*, *Table 1*. Results of multivariate GLM analyses which examines the associations between the principal components (PCs) extracted from 7 CANTAB domains and diagnosis.**

| **Test type** | **Dependent Variables** | **Explanatory variables** | **F** | **df** | **P** | **Partial Eta Squared** |
|---|---|---|---|---|---|---|
| **Multivariate** | All 7 PCs | Diagnosis | 3.34 | 14/206 | <0.001 | 0.175 |
| | | Gender | 1.24 | 7/103 | 0.287 | 0.078 |
| | | Education | 7.16 | 7/103 | <0.001 | 0.327 |
| | | Age | 5.19 | 7/103 | <0.001 | 0.261 |
| Tests for between-subject effects | PC MOT | Diagnosis | 1.51 | 2/109 | 0.226 | 0.027 |
| | PC PAL | Diagnosis | 8.71 | 2/109 | <0.001 | 0.138 |
| | PC RVI | Diagnosis | 6.94 | 2/109 | 0.001 | 0.113 |
| | PC SWM | Diagnosis | 17.34 | 2/109 | <0.001 | 0.241 |
| | PC OTS | Diagnosis | 14.43 | 2/109 | <0.001 | 0.209 |
| | PC IED | Diagnosis | 5.08 | 2/109 | 0.008 | 0.085 |
| | PC ERT | Diagnosis | 11.50 | 2/109 | <0.001 | 0.174 |

Diagnosis: 3 groups, namely healthy controls and schizophrenia with and without the deficit syndrome.

PC MOT: Motor screening test to screen visual, movement and comprehension; PC PAL: Paired-association learning to assess visual memory, episodic memory and learning; PC RVP: Rapid visual information process test to assess visual sustained attention; PC SWM: Spatial working memory to assess working memory and strategy use; PC OTS: One touch stockings of Cambridge to assess spatial planning; PC IED:

Intra/extradimensional set shifting to assess rule acquisition and attention set shifting; PC ERT: Emotional recognition test to interpret facial expression of emotion.

***ESF 3, Tables 1 and 2*** show that nondeficit patients have significant impairments in PC PAL, PC SWM, PC OTS, PC IED, and PC ERT as compared with healthy controls. Deficit schizophrenia was characterized by significant impairments in PC RVP, which were not present in nondeficit schizophrenia, and significantly more dysfunctions in PC PAL, PC SWM and PC ERT, as compared with nondeficit schizophrenia.

***ESF 3, Table 2*. Model-generated estimated marginal means (SE) of the principal components (PCs) extracted from 7 CANTAB domains, in normal controls and schizophrenia patient with and without deficit schizophrenia (SCZ)**

| CANTAB | Normal Control $^{A}$ | Nondeficit SCZ $^{B}$ | Deficit SCZ $^{C}$ |
|---|---|---|---|
| PC MOT | -0.21 (0.13) | -0.13 (0.12) | 0.10 (0.13) |
| PC PAL | -0.46 (0.19) $^{B,C}$ | -0.05 (0.13) $^{A,C}$ | 0.38 (0.19) $^{A,B}$ |
| PC RVP | 0.38 (0.13) $^{C}$ | 0.06 (0.13) $^{C}$ | -0.33 (0.13) $^{A,B}$ |
| PC SWM | -0.62 (0.13) $^{B,C}$ | 0.09 (0.12) $^{A,C}$ | 0.47 (0.13) $^{A,B}$ |
| PC OTS | -0.57 (0.13) $^{B,C}$ | 0.10 (0.12) $^{A}$ | 0.38 (0.13) $^{A}$ |
| PC IED | -0.41 (0.14) $^{B,C}$ | 0.12 (0.13) $^{A}$ | 0.15 (0.14) $^{A}$ |

| | | | |
|---|---|---|---|
| PC ERT | 0.47 (0.12) [B,C] | -0.03 (0.12) [A,C] | -0.37 (0.12) [A,B] |

All results of GLM analysis after covarying for age, sex, and education. [A,B,C]: pairwise comparisons among treatment means.

PC: first principal component extracted from various CANTAB domain tests; PC MOT: Motor screening test to screen visual, movement and comprehension; PC PAL: Paired-association learning to assess visual memory, episodic memory and learning; PC RVP: Rapid visual information process test to assess visual sustained attention; PC SWM: Spatial working memory to assess working memory and strategy use; PC OTS: One touch stockings of Cambridge to assess spatial planning; PC IED: Intra/extradimensional set shifting to assess rule acquisition and attention set shifting; PC ERT: Emotional recognition test to interpret facial expression of emotion.

***ESF 3, Table 3*. Results of multivariate GLM analysis examining the associations between 11 paired-association learning (PAL) test scores and diagnosis [entered as 3 groups: normal controls versus nondeficit schizophrenia (SCZ) versus deficit schizophrenia] while controlling for age, gender and education.**

| **Measurements** | **Controls [A]** | **Nondeficit SCZ [B]** | **Deficit SCZ [C]** | **F** | **df** | **P** |
|---|---|---|---|---|---|---|
| PAL-FTMS | 17.3 (0.8) [B,C] | 14.6 (0.8) [A,C] | 11.2 (0.8) [A,B] | 14.00 | 2/113 | <0.001 |
| PAL-METS | 2.8 (0.5) [B,C] | 4.3 (0.5) [A] | 5.5 (0.5) [A] | 6.28 | 2/113 | 0.003 |
| PAL-MTTS | 2.1 (0.2) [C] | 2.5 (0.2) | 3.1 (0.2) [A] | 4.90 | 3/113 | 0.009 |
| PAL-NPR | 7.5 (0.2) [C] | 7.4 (0.2) [C] | 6.6 (0.2) [A,B] | 4.15 | 2/113 | 0.018 |
| PAL-NPS | 7.1 (0.4) [C] | 6.8 (0.3) [C] | 5.8 (0.3) [A,B] | 3.96 | 2/113 | 0.022 |
| PAL-SC | 7.5 (0.2) [C] | 7.3 (0.2) | 6.7 (0.2) [A] | 3.52 | 2/113 | 0.033 |
| PAL-SCFT | 5.5 (0.2) [B,C] | 4.7 (0.2) [A,C] | 3.7 (0.2) [A,B] | 13.16 | 2/113 | 0.001 |
| PAL-TE | 18.3 (3.3) [B,C] | 28.1 (3.1) [A] | 33.4 (3.2) [A] | 5.41 | 2/113 | 0.006 |
| PAL-TT | 14.1 (0.9) [C] | 16.4 (0.8) | 18.3 (0.8) [A] | 6.28 | 2/113 | 0.003 |
| PAL-TTA | 17.0 (1.9) [C] | 19.8 (1.8) [C] | 26.7 (1.8) [A,B] | 7.18 | 2/113 | 0.001 |

All results are shown as estimated marginal means (SE) obtained after multivariate GLM analysis. F values: results of tests for between-subject effects performed when the multivariate analysis was significant.

[A,B,C]: pairwise comparisons among treatment means. [A]: significantly different from controls, [B:] from nondeficit schizophrenia, [C:] from deficit schizophrenia (protected post-hoc tests)

11 PAL variables are measured, i.e. PAL first trial memory score (FTMS), PAL mean errors to success (METS), PAL mean trials to success (MTTS), PAL number of patterns reached (NPR), PAL number of patterns succeeded (NPS), PAL stages completed (SC), PAL stages completed on first trial (SCFT), PAL total errors (TE), PAL total errors adjusted (TEA), PAL total trials (TT) and PAL total trial adjusted (TTA).

***ESF 3, Table 3*** shows a significant association between diagnosis and the PAL performance (F=2.00, df=22/208, p=0.007) after controlling for age (F=2.06, df=11/103, p=0.030), sex (F=1.91, df=11/103, p=0.047) and education (F=2.07, df=11/103, p=0.029). Table 3 shows the estimated marginal mean (SE) values of the 11 PAL measurements and the tests of between-subject effects for each of the measurements. Tests for between-subject effects showed significant effects of diagnosis on all 11 PAL variables. PAL-FTMS and PAL-SCFT scores were significantly different between the three groups with increasing impairments from controls to nondeficit to deficit schizophrenia. The performance assessed by PAL-NPR, PAL-NPS, PAL-TEA, and PAL-TTA were significantly worse in deficit schizophrenia than in controls and nondeficit schizophrenia. PAL-MTTS, PAL-SC and PAL-TT scores were significantly poorer in deficit schizophrenia versus controls, while results in nondeficit schizophrenia occupied an intermediate position. PAL-METS and PAL-TE were significantly more affected in schizophrenia than in controls.

***ESF 3, Table 4.*** Results of multivariate GLM analyses with the 6 One touch of stocking of Cambridge (OTS) scores examining the associations between 11 Rapid Visual Information Processing (RVP) and diagnosis [entered as 3 groups: normal controls versus nondeficit schizophrenia (SCZ) versus deficit schizophrenia] while controlling for age, gender and education.

| **Measurements** | **Controls** [A] | **Nondeficit SCZ** [B] | **Deficit SCZ** [C] | **F** | **df** | **P** |
|---|---|---|---|---|---|---|
| OTS-MLTC | 25439 (1939) [C] | 20945 (1838) | 18469 (1869) [A] | 3.33 | 2/113 | 0.040 |
| OTS-MLTFC | 16849 (1555) | 14365 (1474) | 12644 (1498) | 1.86 | 2/113 | 0.161 |
| OTS-MCTC | 2.1 (0.1) [B,C] | 2.6 (1.1) [A] | 2.8 (1.1) [A] | 7.65 | 2/113 | 0.001 |
| OTS-PEGC | 0.42 (0.05) [B,C] | 0.69 (0.04) [A] | 0.70 (0.04) [A] | 12.72 | 2/113 | <0.001 |
| OTS-PEGE | 0.36 (0.04) [B,C] | 0.59 (0.04) [A] | 0.64 (0.04) [A] | 15.04 | 2/113 | <0.001 |
| RVP-PFA | 0.008 (0.007) [C] | 0.013 (0.006) [C] | 0.032 (0.006) [A,B] | 3.71 | 2/111 | 0.027 |
| RVP-PH | 0.88 (0.03) [C] | 0.82 (0.03) [C] | 0.72 (0.02) [A,B] | 7.02 | 2/111 | 0.001 |
| RVP-PHB1-7 | 0.88 (0.02) [C] | 0.83 (0.02) | 0.77 (0.02) [A] | 5.00 | 2/111 | 0.008 |
| RVP-TCR | 526.7 (5.6) [C] | 515.7 (5.2) [C] | 493.0 (5.4) [A,B] | 9.89 | 2/111 | <0.001 |
| RVP-TFA | 4.0 (3.3) [C] | 6.5 (3.0) [C] | 15.7 (3.1) [A,B] | 3.80 | 2/111 | 0.025 |
| RVP-TH | 47.4 (1.7) [C] | 44.3 (1.6) [C] | 38.6 (1.6) [A,B] | 7.02 | 2/111 | 0.001 |
| RVP-THB1-7 | 47.4 (1.3) [C] | 44.8 (1.2) | 41.6 (1.3) [A] | 4.50 | 2/111 | 0.008 |
| RVP-TM | 6.6 (1.7) [C] | 9.7 (1.6) [C] | 15.3 (1.6) [A,B] | 7.02 | 2/111 | 0.001 |

| | | | | | | |
|---|---|---|---|---|---|---|
| RVP-TMB1-7 | 6.6 (1.3) [C] | 9.2 (1.2) | 12.4 (1.3) [A] | 4.50 | 2/111 | 0.008 |

All results are shown as estimated marginal means obtained after multivariate GLM analysis. F values: results of tests for between-subject effects performed when the multivariate analysis is significant.[A,B,C]: pairwise comparisons among treatment means. [A]: significantly different from controls, [B:] from nondeficit schizophrenia, [C:] from deficit schizophrenia (protected post-hoc tests)

One Touch Stockings of Cambridge (OTS); OTS mean latency to correct (MLTC), OTS mean latency to first choice (MLTFC), OTS mean choices to correct (MCTC), OTS probability of error given correct (PEGC), and OTS probability of error given error (PEGE).

Rapid visual information process test (RVP) variables, RVP probability of false alarm (PFA), RVP probability of hit (PH), RVP probability of hit blocks 1-7 (PHB1-7), RVP total correct rejections (TCR), RVP total false alarms (TFA), RVP total hits (TH), RVP total hits blocks 1-7 (THB1-7), RVP total misses (TM), RVP total misses blocks 1-7 (TMB1-7).

***ESF 3, Table 4*** shows that there was a significant association between diagnosis and the 6 OTS (F=4.49, df=12/216, p<0.001) scores after controlling for the effects of gender, education, and age (results of multivariate GLM). OTS-MLTC was significantly lower in deficit schizophrenia than in controls, while nondeficit patients took up an intermediate position. OTS-MCTC, OTC-PEGC, and OTS-PEGE are higher in both schizophrenia subgroups as compared with controls. ***ESF, Table 4*** shows that there was a significant association between the diagnosis and RVP values (F=2.70, df=14/210, p=0.001; results of multivariate GLM). Tests for between-subject effects showed significant associations between the diagnosis and all RVP measurements (RVP-A and RVP_ML are shown in Tables 2-3 of the main text). RVP-PFA, RVP-PH, RVP-TCR, RVP-TFA, RVP-TH, and RVP-TM are significantly more disordered in deficit schizophrenia as compared with nondeficit schizophrenia and controls. RVP-PHB1-7, RVP-THB1-7, and RVP-TMB1-7 were significantly more affected in deficit schizophrenia than in controls, while patients with nondeficit schizophrenia occupied an intermediate position.

***ESF 3, Table 5.*** Results of multivariate GLM analysis with the 10 Spatial Working Memory (SWM), 10 Intra-Extra Dimensional Set Shift (IED) and 3 Emotion Recognition Task (ERT) CANTAB measurements as dependent variables and diagnosis as primary explanatory variable [entered as 3 groups: normal controls versus nondeficit schizophrenia (SCZ) versus deficit schizophrenia] while controlling for age, gender and education.

| **Measurements** | **Controls** [A] | **Nondeficit SCZ** [B] | **Deficit SCZ** [C] | **F** | **df** | **p** |
|---|---|---|---|---|---|---|
| SWM-BE4B | 2.2 (0.6) [B,C] | 4.2 (0.6) [A] | 5.4 (0.6) [A] | 5.81 | 2/113 | 0.004 |
| SWM-STR4-10B | 44.0 (1.0) [B,C] | 49.3 (0.9) [A] | 51.1 (0.9) [A] | 14.96 | 2/113 | <0.001 |
| SWM-MTFR | 2420 (302) [C] | 2756 (286) | 3348 (291) [A] | 2.49 | 2/113 | 0.088 |
| SWM-MTLR | 31521 (1977) [C] | 34740 (1871) | 39775 (1905) [A] | 4.53 | 2/113 | 0.013 |
| SWM-MTPT | 1444 (126) [C] | 1573 (119) | 1848 (122) [A] | 2.76 | 2/113 | 0.068 |
| SWM-TE | 32.7 (3.3) [B,C] | 51.6 (3.1) [A,C] | 60.4 (3.2) [B,C] | 18.65 | 2/113 | <0.001 |
| SWM-TE4B | 2.7 (0.8) [C] | 4.2 (0.7) | 6.0 (0.7) [A] | 4.74 | 2/113 | 0.011 |
| SWM-TE4-10B | 32.7 (3.3) [B,C] | 51.6 (3.1) [A,C] | 60.4 (3.2) [B,C] | 18.65 | 2/113 | <0.001 |
| IED-CSE | 8.8 (1.7) [B,C] | 16.0 (1.7) [A] | 14.1 (1.7) [A] | 4.47 | 2/112 | 0.014 |
| IED-CST | 60.0 (4.2) | 71.3 (4.0) | 66.9 (4.1) | 3.05 | 2/112 | 0.052 |
| IED-EB1 | 1.14 (0.66) | 2.53 (0.63) | 2.60 (0.64) | 1.55 | 2/112 | 0.218 |
| IED-PEE | 8.5 (2.3) | 15.7 (2.2) | 13.4 (2.2) | 2.61 | 2/112 | 0.078 |
| IED-SC | 7.4 (0.3) | 7.4 (0.3) | 6.9 (0.3) | 1.02 | 2/112 | 0.365 |
| IED-TE | 25.2 (2.1) [B,C] | 35.9 (2.0) [A] | 36.2 (2.1) [A] | 8.44 | 2/112 | <0.001 |

| | | | | | | |
|---|---|---|---|---|---|---|
| IED-TL | 133042 (18701) [C] | 172280 (17718) [C] | 228994 (18232) [A,B] | 6.76 | 2/112 | 0.002 |
| IED-TT | 85.2 (4.0) [B,C] | 104.7 (3.7) [A] | 108.8 (3.9) [A] | 10.14 | 2/112 | <0.001 |
| IED-TTA | 135.9 (11.7) [C] | 153.8 (11.1) | 170.6 (11.4) [A] | 2.21 | 2/112 | 0.115 |
| ERT-PC | 48.3( 1.8) [B,C] | 41.6 (1.7) [A] | 37.8 (1.8) [A] | 8.34 | 2/114 | <0.001 |
| ERT-TNC | 86.9 (3.3) [B,C] | 74.9 (3.3) [A] | 68.1 (3.2) [A] | 8.34 | 2/114 | <0.001 |

All results are shown as estimated marginal means obtained after multivariate GLM analysis. F values: results of tests for between-subject effects performed when the multivariate analysis is significant [A,B,C]: pairwise comparisons among treatment means. [A]: significantly different from controls, [B:] from nondeficit schizophrenia, [C:] from deficit schizophrenia (protected post-hoc tests)

SWM between errors 4 boxes (BE4B), SWM between errors 4-10 boxes (BE4-10B), SWM strategy 4-10 boxes (STR4-10B), SWM mean time to first response (MTFR), SWM mean time to last response (MTLR), SWM mean token time preparation time (MTTPT), SWM total errors (TE), SWM total error 4 boxes (TE4B) and SWM total errors 4-10 boxes (TE4-10B).

IED completed stage errors (CSE), IED complete stage trials (CST), IED errors block 1 (EB1), IED Pre-ED errors (PEE), IED stages completed (SC), IED total errors (TE), IED total latency (TL), IED total trials (TT) and IED total trials adjusted (TTA).

ERT total trials adjusted (TTA), ERT total number correct (TNC).

***ESF, Table 5*** shows that there was a significant association between the diagnosis and 9 SWM measurements (F=2.97, df=18/210, p<0.001, results of multivariate GLM) after controlling for age, sex and education (results on SWM_BE and SWM_STR are shown in Tables 2-3 of the main text). Tests for between-subject effects showed that SWM-BE4B, and SWM-STR4-10B were significantly more negatively affected in both schizophrenia groups than in controls. SWM-MTFR, SWM-MTLR, SWM-MTPT and SWM-TE4B showed a significantly worse performance of deficit schizophrenia patients than of controls. SWM-TE was significantly different between the three study groups and increased from controls to nondeficit schizophrenia to deficit schizophrenia.

***ESF, Table 5*** shows that there was a significant association between the diagnosis and the 10 IED measurements (F=2.45, df=16/210, p=0.002, results of multivariate GLM analysis; results on IED_TEA and IED_EDS are shown in tables 2-3 of the main text). IED-CSE, IED-TE, and IED-TT were significantly higher, showing worse performance, in both schizophrenic groups than in controls. IED total latency -TL was significantly greater in deficit schizophrenia as compared with controls and nondeficit schizophrenia. IED-TTA was significantly greater (worse result) in deficit schizophrenia than in controls, whereas patients with nondeficit schizophrenia took up an intermediate position.

***ESF, Table 5*** shows that there was a highly significant association between the diagnosis and 3 ERT measurements (F=6.84, df=4/226, p<0.001). Table 2-3 in the main text showed that ERT median overall response latency (MORL) was significantly different between the three groups and increased from controls to nondeficit to deficit schizophrenia. ERT-PC and ERT-TNC (correct results) were significantly lower in both patient groups than in controls.

***ESF 3, Table 6***. Results of 2 different automatic stepwise logistic regression analyses with 1) diagnosis of schizophrenia (SCZ) and 2) deficit schizophrenia (versus non-deficit schizophrenia) as dependent variables.

| **Dichotomies** | **Explanatory variables** | **Wald** | **df** | **p** | **Odds ratio** | **CI 95%** |
|---|---|---|---|---|---|---|
| #1a SCZ vs HC | RVP_ML | 13.81 | 1 | <0.001 | 1.05 | 1.02 – 1.07 |
| | SWM_IED | 8.26 | 1 | 0.004 | 1.05 | 1.01 – 1.08 |
| | ERT-MORL | 8.15 | 1 | 0.004 | 1.001 | 1.00 – 1.001 |
| | Male gender | 6.65 | 1 | 0.010 | 0.20 | 0.06 – 0.68 |
| #1b | RVP_ML | 5.89 | 1 | 0.015 | 3.28 | 1.26 – 8.54 |
| | VFT | 13.84 | 1 | <0.001 | 0.83 | 0.75 – 0.92 |
| | WLM | 5.52 | 1 | 0.019 | 0.85 | 0.75 – 0.96 |
| #2a DEFICIT vs NON | PAL-SCFT | 5.28 | 1 | 0.022 | 0.69 | 0.50 – 0.95 |
| | SWM-STR | 4.79 | 1 | 0.029 | 1.12 | 1.01 – 1.23 |
| | ERT-MORL | 4.49 | 1 | 0.034 | 1.00 | 1.00 – 1.01 |
| #2b. | True Recall | 10.17 | 1 | 0.001 | 0.63 | 0.47 – 0.84 |

CI 95%: 95% confidence interval, lower - upper limit. SCZ vs HC: results of automatic stepwise logistic regression with schizophrenia as dependent variable and no schizophrenia as reference group. DEFICIT vs NON: results of automatic stepwise logistic regression with deficit schizophrenia as dependent variable and no deficit schizophrenia (controls and nondeficit schizophrenia) as reference group

SWM-BE: Spatial Working Memory, between errors; IED-TT: Intra-Extra Dimensional Set Shift, Total Trials; ERT-MORL: Emotion Recognition Task,  mean overall response latency; PAL-SCFT: Paired association learning, stages completed on first trial; SWM-STR: Spatial Working Memory, strategy.

***ESF 3, Table 6*** shows the results of two different automatic stepwise logistic regression analyses with diagnosis as dependent variable and the CANTAB measurements as explanatory variables. As explanatory variables, we selected the three most significant PAL, OTS, RVP, SWM and IED scores and ERT-MORL. Regression # 1a shows that gender and three CANTAB variables were significantly associated with schizophrenia, namely SWM-BE, IED-TT and ERT-MORL ($X^2$= 63.76, df=4, p<0.001, Nagelkerke=0.586, 81.0% of all cases were correctly classified with a sensitivity of 77.9% and a specificity of 87.2%). In regression #1b we examined the 9 key CANTAB tests combined with the CERAD tests and MMSE and found that WLM, VFT and RVP_ML were the most significant predictors ($X^2$= 89.73, df=3, p<0.001, Nagelkerke=0.548, 82.8% of all cases were correctly classified with a sensitivity of 88.3% and a specificity of 71.8%).

Regression #2a shows the outcome of a second logistic regression analysis with deficit schizophrenia as the dependent variable and nondeficit schizophrenia as the reference group. We found that SWM-STR and ERT-MORL were significantly and positively associated with deficit schizophrenia whereas PAL-SCFT was negatively related to deficit schizophrenia ($X^2$=30.69, df=3, p<0.001, Nagelkerke=0.326, 77.6% of all cases were correctly classified with a sensitivity of 70.3% and a specificity of 81.0%). In regression #2b we examined the 9 key CANTAB tests combined with the CERAD tests and MMSE and found that True Recall was the only variable discriminating deficit from nondeficit schizophrenia ($X^2$=12.81, df=1, p<0.001, Nagelkerke=0.204, 67.5% of all cases were correctly classified with a sensitivity of 64.9% and a specificity of 70.0%).

***ESF 3, Discussion***

Moreover, some tests, which are not considered to be key tests by CANTAB [12] are more relevant for deficit schizophrenia than the key tests of the same domain. For example, PAL_FTMS, IED_TE, IED_TT, RVP_TCR, RVP_TM, and RVP_TH are more significantly associated with deficit schizophrenia than the key tests in the same domain, namely PAL_TEA, IED_TEA, and RVP_A. In the discrimination of deficit from nondeficit schizophrenia we found that PAL_SCFT, which is not a key test according to CANTAB, has more discriminatory power than the PAL key tests. When discriminating schizophrenia from controls, SWM_IED is a more significant discriminatory test than the key CANTAB tests. Furthermore, using the first PC extracted from all tests in the subdomains allowed us to detect additional aberrations in the SWM and IED domains in deficit schizophrenia.
